# Pathogenic Neurofibromatosis type 1 (NF1) RNA splicing resolved by targeted RNAseq

**DOI:** 10.1101/2021.04.22.21252609

**Authors:** R. Koster, R.D. Brandão, D. Tserpelis, C.E.P. van Roozendaal, C.N. van Oosterhoud, K.B.M. Claes, A.D.C. Paulussen, M. Sinnema, M. Vreeburg, V. van der Schoot, C.T.R.M. Stumpel, M.P.G. Broen, L. Spruijt, M.C.J. Jongmans, S.A.J. Lesnik Oberstein, A.S. Plomp, M. Misra-Isrie, F.A. Duijkers, M.J. Louwers, R. Szklarczyk, K.W.J. Derks, H.G. Brunner, A. van den Wijngaard, M. van Geel, M.J. Blok

## Abstract

**Purpose:** Neurofibromatosis type 1 (NF1) is caused by loss-of-function variants in the *NF1* gene. Approximately 10% of these variants affect RNA splicing and are either missed by conventional DNA diagnostics or are misinterpreted by *in silico* splicing predictions. A targeted RNAseq-based approach was designed to detect pathogenic RNA splicing and associated pathogenic DNA variants.

**Methods:** RNA was extracted from lymphocytes, followed by targeted *NF1* RNAseq. An in-house developed tool (QURNAS) was used to calculate the enrichment score (ERS) for each splicing event.

**Results:** This method was thoroughly tested using two different patient cohorts with known pathogenic splice-variants. In both cohorts all 56 normal reference transcript exon splice junctions, 24 previously described and 45 novel non-reference splicing events were detected. Additionally, all expected pathogenic splice-variants were detected. Eleven patients with NF1 symptoms were subsequently tested, three of which have a known *NF1* DNA variant with a putative effect on RNA splicing. This effect could be confirmed for all 3. The other eight patients were previously without any molecular confirmation of their NF1-diagnosis. A deep-intronic pathogenic splice variant could now be identified for two of them (25%).

**Conclusion:** Targeted *NF1* RNAseq can be successfully used to detect pathogenic RNA splicing variants, complementary to DNA based diagnostics.

## Introduction

Neurofibromatosis type 1 (NF1; MIM 162200) is one of the most common autosomal dominant tumor-predisposition disorders affecting ∼1 in 3000 individuals.^1,2^ Patients typically present with the following clinical manifestations: café-au-lait spots, dermal neurofibromas, iris hamartomas (Lisch nodules), axillary and/or inguinal freckling, and subcutaneous or plexiform neurofibromas. Other manifestations include learning disabilities, optic glioma, skeletal abnormalities and an increased risk of specific malignancies.^1-3^ Malignancies are an important cause of the 8–15-year reduction in average life expectancy.^1-3^ Most patients are clinically diagnosed in childhood, according to NIH consensus criteria.^4^ However, genetic testing to confirm a clinical diagnosis is still warranted, because of the clinical overlap with Legius syndrome (MIM 611431; *SPRED1)*.^1,2,5^ In addition, an accurate genetic diagnosis facilitates appropriate screening and follow-up, reproductive options (prenatal testing and pre-implantation genetic diagnosis) and access to clinical studies and potential future trials and treatments.

NF1 is caused by loss-of-function variants in the tumor-suppressor gene *NF1* (MIM 613113), located on chromosome 17q11.2 and consisting of ∼350 kb of genomic DNA. The *NF1* gene produces a major 12kb transcript NM_000267.3 that contains 57 exons and encodes for a Ras-guanosine triphosphatase (GTPase) activating protein: neurofibromin.^1,6-8^ Pathogenic variants in *NF1* result in loss of function of neurofibromin causing an increase in Ras signaling, affecting cell proliferation and differentiation.^9^

Currently, there are almost 3700 (likely) pathogenic *NF1* variants reported in HGMD (HGMD® Professional 2020.4) and 1890 in Clinvar (Jan 2021),^10,11^ and at least half of these arose *de novo*.^12-16^ The majority of (likely) pathogenic variants in *NF1* are predicted to produce a truncated form of neurofibromin, and of these variants ∼30% cause splicing alterations affecting mRNA processing.^12-16^ At least a third of these splicing alterations are either not found using standard DNA diagnostics (targeting only the coding region, several intronic nucleotides, but not including potential retrotransposon insertions) or are variants not predicted to affect splicing by current bioinformatics algorithms.^12-17^

The diagnostics laboratory community has adopted the ACMG standards and guidelines for the interpretation of sequence variants^18^ to determine their clinical relevance, including recommendations for splice variants. For variants adjacent to exon boundaries and not further than two nucleotides from the exon border, *in silico* tools often adequately predict changes in RNA splicing.^19^ However splice prediction tools for variants outside the consensus splice acceptor/donor sites perform poorly.^20^ And although these tools are able to predict cryptic or novel splice sites, the *in vivo* outcome of competing splice sites for the splicing complex is difficult to predict.^21^ Moreover, the specificity of predictions for the effect of genetic variants on splice enhancer or silencer sites, potential branch points^22^ or even change in the branch point-exon distance^23^ is limited. As a consequence, the effect of DNA variants on RNA splicing ultimately needs to be determined experimentally.

The experimental verification method most often employed for splice variant validation is conventional Reverse Transcription (RT)-PCR, which has its drawbacks. First, it is laborious, time consuming and prone to preferential amplification of transcripts depending on primer choice. Another possible pitfall is the use of primers that amplify only a small part of the region surrounding the variant, while the genetic effect may involve skipping of multiple exons.^21^ Furthermore, data are only semi-quantitative and, finally, almost every new splice isoform needs its own RT-PCR primer design. Targeted RNAseq on the other hand gives comprehensive information about RNA expression and splicing for genes/transcripts and can circumvent the aforementioned draw-backs.

A diagnostic procedure was designed that uses an in-house developed tool QURNAs (**Qu**antitative enrichment of aberrant splicing events in targeted **RNAs**eq) to facilitate the detection and quantification of normal and/or pathogenic RNA splicing events in targeted *NF1* RNA sequencing data.

## Materials and methods

### Patient derived cells

For the initial validation of the targeted RNAseq procedure, RNA was isolated from uncultured peripheral blood lymphocytes of nine patients from the pre-implantation genetic testing (PGT) program at our institution, who had a pathogenic *NF1* variant that leads to aberrant splicing (Table 1). Two additional samples from anonymous patients without a clinical diagnosis of NF1 were included as wild type controls. These validation samples were enriched for *NF1* RNA using NuGEN SPET for RNA (see RNA isolation and Target enrichment/Library preparation).

**Table 1.** Overview of validation patient samples, including nine samples with a known pathogenic *NF1* splice-variant and two anonymous patients without a clinical NF1 diagnosis. Expected and/or published splicing events are shown together with the top events found using QURNAs. #1,2,4 are artefacts; #3 ΔExon5 natural occurring event (ERS ∼0.5; 731 reads across all samples); #5 Δ15 natural occurring event (ERS ∼0.3 ERS; 144 reads across all samples). ^ FS = frame shift; IF = in frame. ^*1^Wimmer et al. 2007^13^; ^*2^Pros et al. 2008^31^; ^*3^De Conti et al. 2012^32^; ^*4^Messiaen et al. 1999^34^

| Sample | NF1 variant<br>(NM_00267.3) | Expected/published<br>splicing events^ | Top splicing event (effect)^ | Top event ERS (reads) | RNA | Protein |
| --- | --- | --- | --- | --- | --- | --- |
| Val. 01 | c.7127-1G>A | Δ1nt 5'exon 48 (FS) | Δ1nt 5'exon 48 (FS) | 16.5 (999) | r.7128del | p.(Tyr2377Thrfs*20) |
| Val. 02 | c.654+2insT | Δexon 6 (FS) | Δexon 6 (FS)<br>Δexon 6,7 (IF) | 23.5 (2676)<br>8.1 (1063) | r.587_654del<br>r.587_730del | p.(Glu196Glyfs*12)<br>p.(Thr197_Glu244del) |
| Val. 03 | c.4773-2A>G | use of cryptic acceptor site,<br>Δ293nt 5'exon 36 (FS)*<sup>1,2</sup> | Δ293nt 5'exon 36 (FS)<br>+multiple (incl Δ36) | 15.1 (392)<br>range 6.3-10.2 (295-400) | r.4773_5065del<br>multiple | p.(Phe1592Leufs*7)<br>Δ36=p.(Phe1592Leufs*8) |
| Val. 04 | WT | - | Δ5' part exon 37 - 5' part exon 41 (IF) | 5 (150)<sup>#1</sup> | r.4986_6506del | p.(Trp1664_Ser2170del) |
| Val. 05 | WT | - | splicing 3'UTR | 6.7 (90)<sup>#2</sup> | r.? | p.? |
| Val. 06 | c.586+5G>A | Δexon 5 (FS)<sup>*3</sup> | Δexon 5 (FS) | 6.1 (6506)<sup>#3</sup> | r.480_586del | p.(Leu161Asnfs*4) |
| Val. 07 | c.1466A>G<br>(p.Tyr489Cys) | Δ62nt 3'exon 13 (FS)<sup>*4</sup> | Δ45nt exon 18 (IF)<br>Δ62nt 3'exon 13 (FS) | 23.4 (228)<sup>#4</sup><br>22.4 (1792) | r.2106_2150del<br>r.1466_1527del | p.(Val703_Ala717del)<br>p.(Tyr489*) |
| Val. 08 | c.4515-15_4515-13delGTT | ins 14nt 5'exon 34 (FS) | ins 14nt 5'exon 34 (FS) | 19.2 (3123) | r.4514_4515ins4515-17_4515-1del4515-15_4515-13 | p.(Arg1505Serfs*53) |
| Val. 09 | c.1721+2T>A | Δexon 15 (FS) | Δexon 15 (FS) | 9.6 (2793)<sup>#5</sup> | r.1642_1721del | p.(Ala548Leufs*13) |
| Val. 10 | c.5205+1G>C | Δ54nt 3'exon 36 (IF) | Δ54nt 3'exon 36 (IF) | 44.9 (9158) | r.5152_5205del | p.(Phe1719_Val1736del) |
| Val. 11 | c.1885G>A<br>(p.Gly629Arg) | Δ41nt 5'exon 17(FS)<sup>*2</sup> | Δ41nt 5'exon 17 (FS)<br>Δ46nt 5'exon 17 (FS) | 18.1 (1420)<br>5.2 (703) | r.1846_1886del<br>r.1846_1891del | p.(Gln616Glyfs*4)<br>p.(Gln616Aspfs*57) |

The diagnostic performance of targeted *NF1* RNAseq was further assessed using samples from the Centre for Medical Genetics, Ghent University Hospital, Ghent, Belgium (Table 2). These blood lymphocytes were short-term cultured in RPMI1640 with 10% FCS supplemented for stimulating growth with IL-2 and PHA^20^, besides puromycin as an inhibitor of nonsense mediated RNA decay (NMD)^24^. The molecular results of these samples were initially blinded, to test the ability to detect pathogenic *NF1* splicing events without prior knowledge, comparable to the diagnostic setting. These replication cohort samples were enriched for *NF1* using both Nugen SPET for RNA and for *NF1*/*SPRED1* using Agilent SureSelect RNA Target Enrichment (see RNA isolation and Target enrichment/Library preparation). This allowed a direct performance comparison of these two different commercial platforms.

**Table 2.**
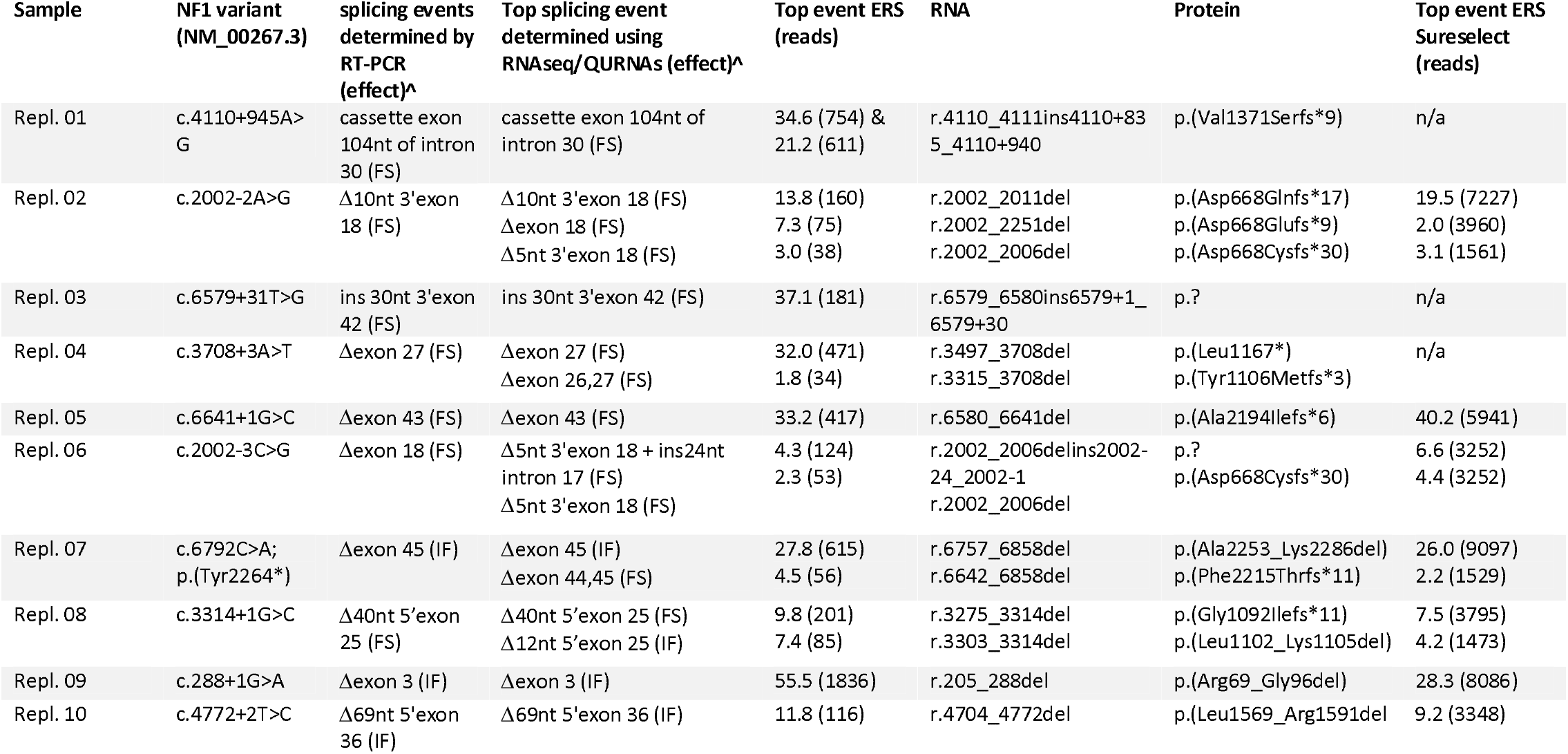
Overview of replication patient samples, including ten samples with a known pathogenic *NF1* splice-variant (initially we were blinded for the variants). The previously determined splicing events using RT-PCR and Sanger sequencing by an external clinical lab are shown, together with the top events found using QURNAs. ^ FS=frame shift; IF=in frame.

To further validate the targeted RNAseq approach and show its added value for the detection of pathogenic splicing events complementary to DNA diagnostics, an additional set of twelve patients was selected (Table 3). Eight patients were included because of having (or suspected of having) a clinical NF1 diagnosis without molecular confirmation. While two patients had a *NF1* DNA variant with unclear effect on RNA splicing (variant of unknown significance – VUS [class 3]) and one patient had an *NF1* DNA variant likely to affect splicing (likely pathogenic – LP [class 4]). One additional sample from an anonymous patient without a clinical diagnosis of NF1 was included. Available material from these patients, i.e. either lymphocytes directly isolated from blood or short term cultured cells with or without NMD inhibition, was used to isolate RNA. These samples were enriched for *NF1*/*SPRED1* using SureSelect RNA Target Enrichment (see RNA isolation and Target enrichment/Library preparation).

**Table 3.**
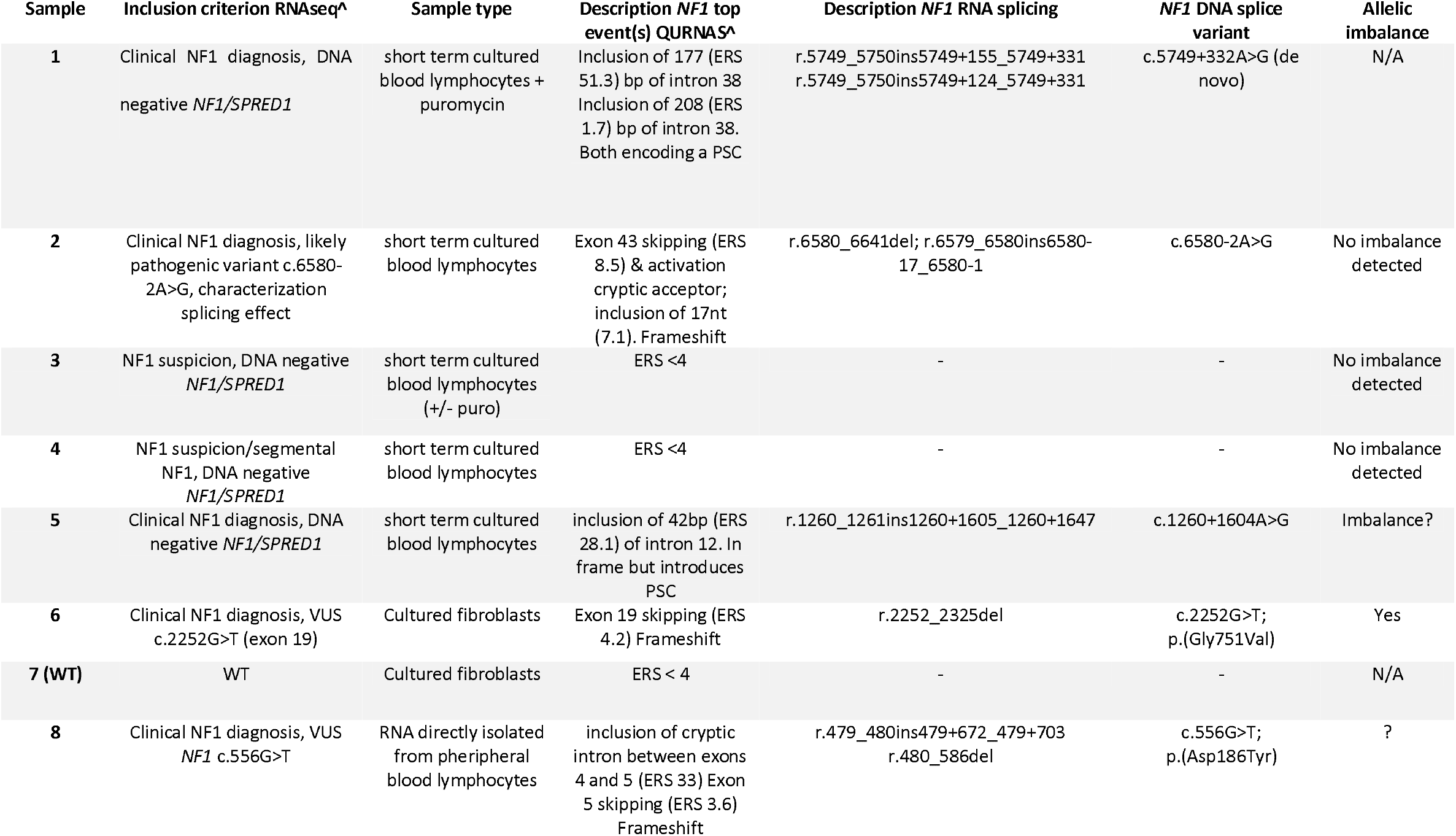
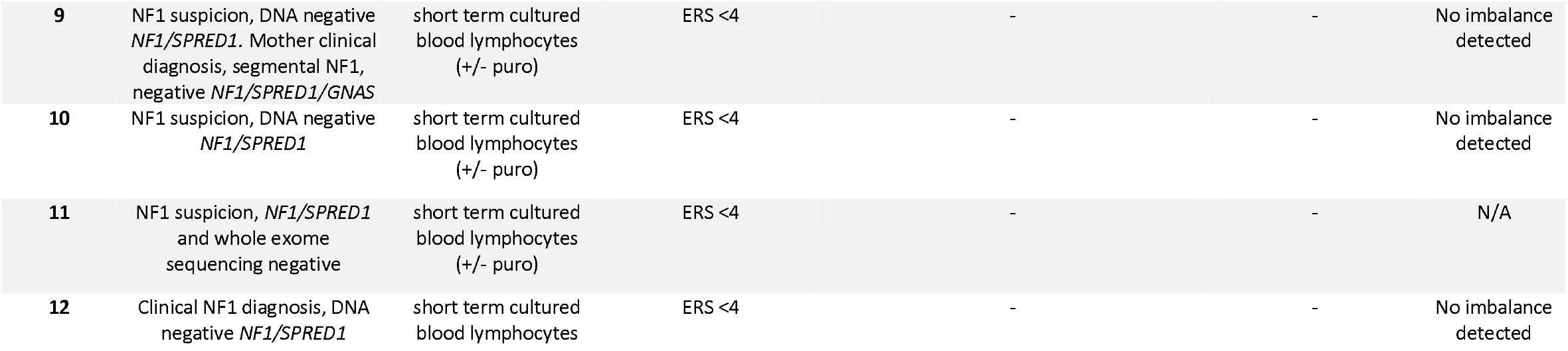
Overview of molecular undiagnosed patients. These were suspected for, or having a clinical NF1 diagnosis without (clear) molecular diagnosis. Inclusion criteria and top event(s) determined with QURNAs are shown. ^LP=likely pathogenic; VUS=variant of unknown significance; PSC=predicted stop codon. Clinical NF1 diagnosis = according to NIH consensus criteria. NF1 suspicion = did not meet minimal NIH consensus criteria.^4^

The samples used in this study were from patients undergoing diagnostic genetic testing. All patients provided written or oral consent for coded use of their material for both improving diagnostic testing and to study clinical utility.

### RNA isolation and Target enrichment/Library preparation

Total RNA was purified and DNAseI treated using RNeasy Plus micro columns (Qiagen) and used as input for the Ovation cDNA module followed by custom Ovation target enrichment, based on single primer enrichment technology (SPET) for RNA, according to the protocol of the manufacturer (NuGEN). Primer design for *NF1* (NM_000267.3) was performed by the manufacturer for the coding regions and 5’/3’UTR with a probe spacing of 150bp. Library preparation was according to the manufacturer instructions.

Additionally, the SureSelect RNA Target Enrichment for Illumina Paired-End Multiplexed Sequencing kit (Agilent; protocol version 2.2.1) was used to target the *NF1 (*NM_000267.3) and *SPRED1* (NM_152592.2) coding regions and 5’/3’UTR. Probe design was performed using the Agilent eArray online tool, setting the region of interest to minus 100nt of the normal 5’exon boundary and plus 150nt of the normal 3’exon boundary and 6x tiling of probes. Library preparation was according to the manufacturer’s instructions. Primers (SPET) and/or probes (Sureselect) design is available upon request.

Paired-end sequencing (2×150bp) was performed on a NextSeq500 instrument (Illumina) using NextSeq500/550 high output kit v2 (Illumina).

### Read alignment

RNAseq reads were mapped with the STAR mapper (Version 2.4.1d) and an index created from HS.GRCh37 with 92bp overhang.^25^ We performed unique mapping onto the genome with max. 10 mismatches per read and no chimeric reads were allowed. To elucidate the PCR amplification bias, de-duplication was performed either using a build in Unique Molecular Identifier (UMI) in the probe design (NuGen SPET) or by removing identical reads (Agilent Sureselect). Duplicate read pair detection was carried out with Picard tools (https://github.com/broadinstitute/picard). Start and end positions from STAR output refer to the first nucleotide in the intron (AG|**<u>g</u>**u) and last nucleotide of the intron (a**<u>g</u>**|G), respectively.

### Quantitative enrichment of aberrant splicing events in targeted RNAseq – QURNAs

The QURNAs computational pipeline was developed to identify sample-specific splicing events in targeted RNAseq data, including novel and possibly aberrant transcripts or increased abundance of normal isoforms (see **Supplemental figure 1**). QURNAs is available at https://dataverse.nl/dataset.xhtml?persistentId=hdl:10411/LY8ZQ4. The mapping of short sequences to the reference genome without prior assumptions about transcript architecture, enables statistical evaluation of significance of observed RNA isoforms.

### *De novo* prediction of splice junctions from RNAseq data

QURNAs first creates a collection of splice sites based on the reads mapped onto the genome (BAM file). For these splice sites, reads in BAM files that overlap with the splice site are counted. While iterating over all reads in the region, two additional features are counted to estimate the fraction of the reads in the region congruent with the splice junction. First, the number of reads is calculated that overlap with and contain the last nucleotide of the exon at the donor site. This includes, next to reads with a different acceptor site, “unbroken” reads corresponding to intron retention events. The second count consists of reads that “span” the splice site, thus start upstream of the nucleotide and end downstream, but do not necessarily contain the nucleotide next to the donor site in their sequence (*e*.*g*., exon skipping events or splice events that use an alternative upstream donor site).

### Enrichment score (ERS) calculation

Enrichment for a splicing event in a sample is calculated as follows. For each splice site, a relative read count *l*^*spl*^ is calculated that measures the fraction of aligned reads that are congruent with the splice site among all the reads that span the splice site. Formally, we count reads (*r*^*spl*^) that are congruent with the splice *spl* = (start,end) *i*.*e*., the genomic alignment of the read contains a gap between exact (start, end) positions, allowing for other insertions/deletions in the read. Reads for each of the splice *r*^*spl*^ are considered relatively to reads that *span* the splice site *s*^*spl*^ (spanning reads). These start their genomic alignment upstream of the donor site and end downstream of this site but do not necessarily share the same donor or acceptor sites. To minimize the impact of spurious reads and sequencing noise, pseudocounts of at least 10 reads and up to 1% of spanning reads (max(10,*s*^*spl*^ ·1%)) are added to both *r*^*spl*^ and *s*^*spl*^. The relative read count *l*^*spl*^ for each sample is then calculated as

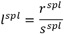

In sample *i*, after calculating the relative splice read count, enrichment of reads 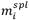 is calculated by comparing the relative read count to the average read counts from other samples

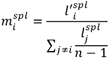

To prevent samples with low sequencing coverage having inflated relative read counts due to pseudocounts (e.g. 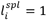 for the extreme case of absent coverage, i.e. 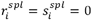), the enrichment 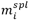 is calculated using 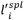 without pseudocounts.

### Statistical significance of enriched isoforms

To test whether splice isoforms are expressed at significantly higher levels in a sample, a two-tailed proportion test that tests the null hypothesis that proportions in several groups are the same is used. For each splice junction a group of trials as number of spanning reads (*s*^*spl*^) that includes reads that support the junction *r*^*spl*^ (positive outcome of the trial) is defined. If significant (*p*(*spl*) < 0.01), the proportion test rejects the null hypothesis that the expression of the splice junction is the same in all samples. The test assumes independence between trials in the group and this is not the case for RNAseq due to read redundancy and overlap between 150-bp reads. The marginal information contributed by additional reads, drops with the number of reads mapped to a splice junction. To reflect lower marginal information, we use square root of RNAseq read numbers:

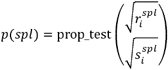

where *i* corresponds to subsequent samples. To determine the significance of a splice event in a specific sample *i*, the reads for the sample are compared to the background distribution of reads in remaining samples, testing the enrichment hypothesis (one-tailed test expecting enrichment):

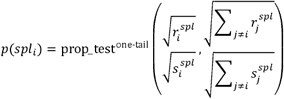

Note that in this statistical framework it is possible that ***p***(*spl*) < 0.01 but ***p***(*spl*_*i*_) > 0.01 for all samples ***i*** (the distribution of reads is different between samples, but none are significant). This is, for example the case for WT splice isoforms in mutated samples where the WT isoform becomes depleted ***p***(*spl*_*i*_) tests only for enrichment).

### Interpretation QURNAS output

The QURNAs output for all splicing events was reduced with an additional GAP filter of >3 ((pos end_STAR – pos start_STAR) +1), to exclude artefacts that do not represent splicing since the intron size would be smaller than 4 nucleotides. The absolute read counts of all normal exon splice-junctions in the reference transcript were inspected to confirm good sequence coverage throughout the gene for all samples. The ERS values of these reference exon splice junctions were subsequently checked for a possible drop (normal reference exon splice junctions are expected to have a median ERS ∼1). Highly expressed alternative splicing events usually result in lower expression of the normal exon splice junctions in the same region, reflected by such a drop. Subsequently, all splicing events for a specific sample were sorted high to low based on the ERS. All events with ERS > 5 were further evaluated to discriminate possible artefacts from true alternative splicing events. Importantly, relevant ERS values can be lower than 5 when; 1) the splice variant gives rise to multiple different splicing events, instead of one or two major events; 2) the splicing events concerns an upregulated normal splicing event which is also present in other samples in the run at lower expression levels or 3) when 2 samples are present in the same run with the same effect on splicing. Alamut v2.15 (Interactive Biosoftware) was used to identify true splice-donor and acceptor sites based on *in silico* prediction. Regions with alternative splicing were further inspected in the BAM file in a genome browser (IGV and Alamut) to check for possible causal sequence variants in close proximity. Allelic imbalance was assessed by comparing heterozygous variants in the DNA vs the RNA sequences in IGV.

### Nomenclature

The description of genetic variants follows the Human Genetic Variation Society (HGVS) approved guidelines,^26^ where c.1 (and r.1) is the A of the ATG translation initiation codon. Alternative splicing events are those incorporating splice junctions not present in the reference transcript NM_000267.

## Results

### QURNAs detects normal and known pathogenic *NF1* splicing events

To validate our diagnostic procedure, targeted *NF1* RNAseq was performed using blood samples of nine patients with a known pathogenic *NF1* splice-variant, and two control samples. (**Table 1**).

First, the presence of all normal exon-exon splice junctions belonging to *NF1* reference transcript NM_000267 was checked and found detectable with a median enrichment score (ERS) ∼1 and >5000 reads per splice junction across all samples (**Figure 1a, Supplemental Table 1**,**2**). Furthermore, with QURNAs an extensive list of (naturally occurring) splicing events for *NF1* was obtained (**Supplemental Table 2)** and compared to previously published naturally occurring events (**Supplemental Table 3)**. From the 25 previously reported normal (rare) events, 24 were detected. Approximately half of these previously reported events are major splicing events, detected by QURNAs in almost all of the patient derived cells (median ERS∼1 & reads >1000). The other half are minor events (median ERS<1 & reads <700) and detected in a fraction of the patient derived cells (**Supplemental Table 2)**. The only missed previously reported normal rare event is from a transcript ‘ins 9a/9br’ (legacy exon nomenclature, corresponding to insertion of 10 amino acids between exons 11-12 of *NF1;* c.1260+1617_1260+1646) that is supposed to be specific for neuronal tissue, and not detectable in blood.^27-29 33^ Additionally 45 events not published before were detected in multiple samples (**Supplemental Table 2**).

**Figure 1.**
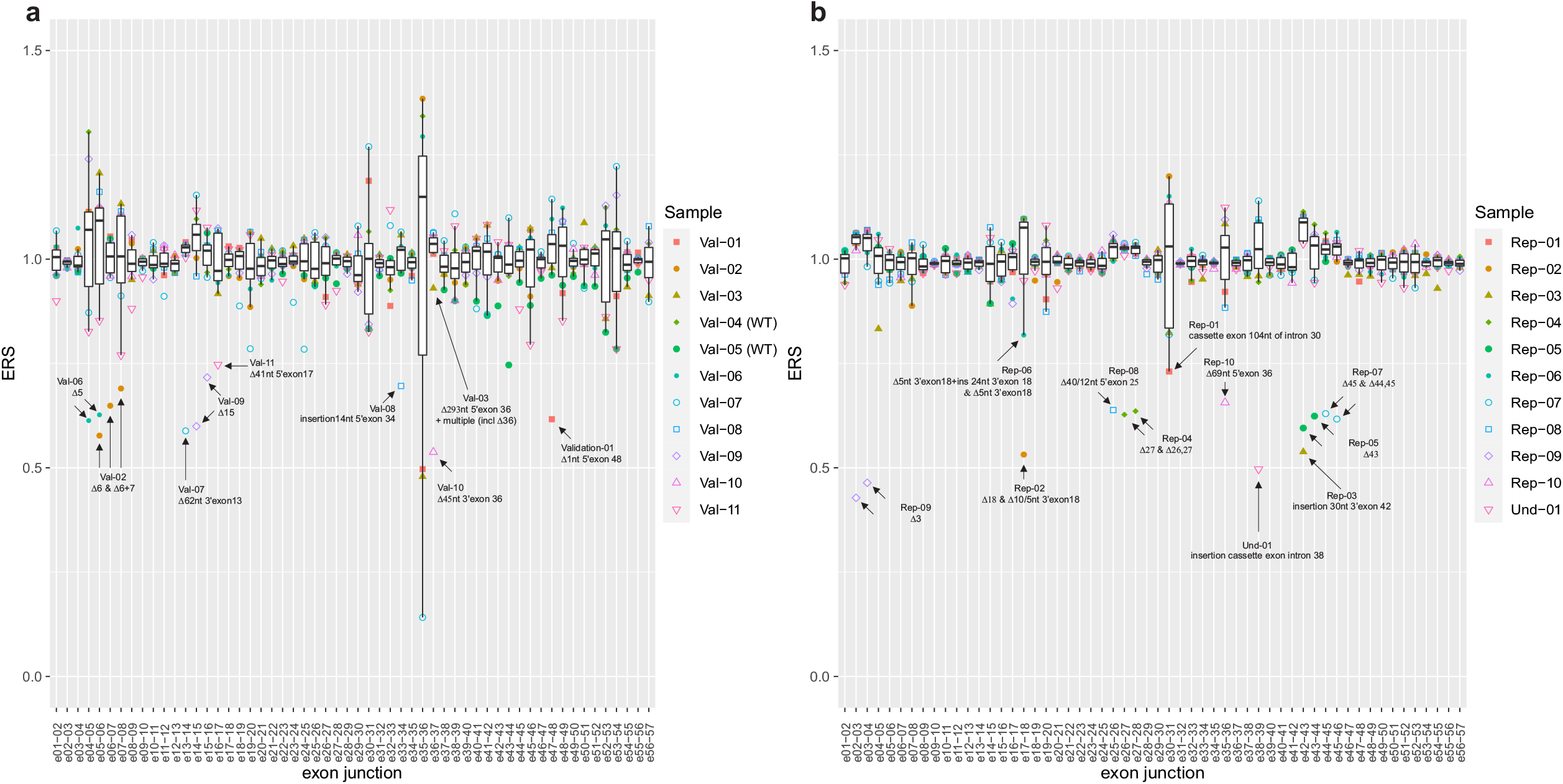
Drop in enrichment score of normal exon splice junctions for all NF1 patients is clearly visible in close proximity to their specific pathogenic splicing event. Enrichment score of normal exon-exon splice junctions of the validation cohort (a) and the replication cohort (b). Shown are the ERS score for the normal exon splice junctions for the validation (a) and replication (b) cohort. Annotated in the figures are the drop in ERS due to the pathogenic splicing event. Normal reference exon splice junctions are expected to have a median ERS ∼1.

For the nine samples with a known pathogenic splice-variant all expected (major) pathogenic splicing events were detected using QURNAS. The corresponding causal DNA variant could also be observed in the RNAseq reads (data not shown). The highest ERS per sample and concomitant major drop in ERS of the normal exon-exon splice-junctions due to alternative splicing are listed (**Table 1, Supplemental Table 1**,**2 & Figure 1a)**. Samples 2,3 and 11 had additional (minor) splicing events that are related to their major pathogenic splicing event albeit with lower ERS. Events with a maximum ERS of ∼7 were found for the two control samples. However, these events had relatively low reads per event (<150) and do not appear to be true splicing events based on *in silico* predictions (**Supplemental Figure 2)**. Also, the concomitant drop in ERS of the normal exon-exon splice junctions in close proximity was absent, indicating that wild-type splicing was not significantly altered. (**Figure 1a)**. This indicates that different data sources have to be taken into account for correct interpretation of an increased ERS, to be able to discriminate artefacts/non-splicing related events from true alternative splicing events. This is further illustrated by samples 6 and 7.

For sample 6 of the validation cohort with variant c.586+5G>A, previously shown to cause skipping of exon 5,^30-32^ QURNAS indicated an ERS of 6.1 for exon 5 skipping (covered by 6509 reads). This is also reflected in figure 1a by a drop in ERS at the exon 4-5 junction (ERS∼0.61) and 5-6 junction (ERS∼0.63) compared to the other samples in the same run (median ERS ∼1 for exon junction 4-5 and median ERS∼1 for 5-6 across all samples – see **Figure 1a, Supplemental Figure 3**). The ERS of 6.1 for exon 5 skipping is smaller than expected, most likely because exon 5 skipping is a (previously published) naturally occurring event (Δ5 or ‘NF1-ΔE4b’)^33^ that is detected with a median ERS of 0.52 and median 728 reads across all samples (**Supplemental Figure 3**). Sample 7 of the validation cohort harbors variant c.1466A>G (p.(Tyr489Cys)) in *NF1*. This specific variant has been reported many times as a disease-causing splice variant, since RNA studies have shown that it creates a new donor site leading to the deletion of 62nt 3’ of exon 13 (**Supplemental Figure 4)**.^15,30,34^ This is confirmed using RNAseq (high ERS of 22.4 and covered by 1792 reads). The event corresponds to a drop in ERS to 0.59 for sample 7 at the junction of exon 13-14 compared to median ERS∼1 across all samples for this event in the same run. The observed drop in ERS is due to partial loss of normal exon 13 to exon 14 splicing. However, one event with a higher ERS score was also observed for this sample (ERS 23.4; 228 reads - **Table 1, Supplemental Table 1**). Again, this event corresponds to an artefact based on *in silico* predictions (**Supplemental Figure 4**), the relatively low read number and the concomitant drop in ERS of the normal exon-exon splice junctions in close proximity is lacking (**Figure 1a**).

### RNAseq replication of RT-PCR based molecular diagnosis

Ten RNA samples from an external diagnostic laboratory that uses RT-PCR to detect pathogenic *NF1* splicing, were subsequently retested using the targeted RNAseq assay and processed with QURNAs. Initially, the RT-PCR results were blinded to us to test the performance of the *NF1* targeted RNAseq assay in a simulated diagnostic setting. The RT-PCR results could be fully replicated with some minor differences and could even be complemented with splicing events that are more difficult to detect or extract from conventional RT-PCR and Sanger sequencing data (**see Table 2; further discussed below**). Using the targeted RNAseq diagnostic procedure all pathogenic splicing events were detected with high ERS for the aberrant event (**Table 2)** and the concomitant drop in ERS of the normal exon-exon splice junctions in close proximity (**Figure 1b**). For these samples, as for the validation set, we could detect all normal reference exon splice junctions (median ERS∼1, median reads>800) and 24 published naturally occurring splicing events that are detectable in blood derived RNA (**Supplemental Table 2**,**4**). Besides detection of the causal splicing events using QURNAs, the corresponding causal DNA variant could be observed in the RNAseq reads using the BAM-file (data not shown).

A minor discrepancy between the RT-PCR and RNAseq data was observed for replication sample 6. RT-PCR determined skipping of exon 18, while RNAseq in combination with QURNAs showed deletion of the first 5nt in combination with the intronic insertion of 24nt (Δ5nt 3’exon 18 + ins24nt intron 17; ERS 4.3 with 124 reads) and deletion of the first 5nt only (Δ5nt 3’exon 18; ERS 2.3 with 53 reads). Skipping of exon 18 is only observed with ERS 0.5 and 16 reads. After thoroughly reinvestigating the RT-PCR Sanger traces, the forementioned events could be observed in the background. An explanation for this difference may be preferential amplification of the smaller Δexon 18 fragment with RT-PCR.

For sample 2, RNAseq identified besides the previously with RT-PCR detected deletion of the first 10nt of exon 18 (ERS 13.8, reads 160), additional exon 18 skipping (ERS 7.3, reads 75) and deletion of the first 5nt of exon 18 (ERS 3.0, reads 38, see **Table 2 & Supplemental Table 3**). After reevaluating the RT-PCR Sanger traces, these events could also be observed as minor events in the background.

### Targeted enrichment platform comparison

Part of the replication cohort samples was used to test the performance of QURNAs in combination with a different library preparation method to enrich for *NF1* transcripts. PCR based targeting (NuGEN SPET) to enrich for *NF1* was switched to a hybridization based enrichment (Agilent SureSelect). Comparable results were obtained with QURNAs using the different enrichment protocols. All splicing events were present using both methods with some variations in ERS and reads (**Table 2 & Supplemental Figure 5)**.

### Testing molecular undiagnosed cases with a (suspected) clinical diagnosis of NF1

Eleven patients with a (suspected) clinical diagnosis of NF1, who lacked a definitive molecular diagnosis based on DNA diagnostics, were tested using targeted RNAseq (Sureselect, Agilent) and QURNAS data-analysis. All patients were checked for the highest ERS event(s) (**Table 3 and supplemental Table 4**,**5**) and a possible drop in the relative expression of the normal exon junctions (**Figure 1b (patient Und-01), 2a (other patients)**). When possible, heterozygous single nucleotide variants present in the coding DNA sequence were compared with the RNA sequence reads, to detect potential allelic imbalance (**Table 3**). For six patients no enriched pathogenic splicing event in *NF1* nor *SPRED1* (*SPRED1* data not shown) could be detected based on high ERS, nor a prominent drop in ERS of the normal exon-exon junction(s). Also, comparing the DNA and RNA sequencing reads, no evident allelic imbalance was observed for these patients. Interestingly, for five patients we were able to identify a deep intronic pathogenic splice variant or resolve the exact effect on RNA splicing for variants (VUS, likely pathogenic) previously detected using DNA diagnostics (**Table 3**).

For undiagnosed patient 1 (in the same run as the replication samples), two major enriched splicing events between exons 38 and 39 were detected. These events introduce a cassette exon of 177bp in intron 38 (acceptor site: ERS 51.3 and 1170 reads and donor site ERS 20.4 and 273 reads). In addition, a minor alternative acceptor site was also seen in the QURNAS output, resulting in a larger cassette exon of 208bp at the 5’end but having the same 3’end (ERS 1.4 and 31 reads). The inclusion of these novel cassette exons, results in a drop in ERS of the normal 38-39 exon-exon junction (**Figure 1b,2b**). These aberrant splicing events are caused by a deep intronic c.5749+332A>G change, that is visible in the RNAseq reads (**Supplemental Figure 6)**. The presence of this variant was confirmed with Sanger DNA sequencing as a *de novo* pathogenic splice variant by testing the patient and parents. This variant introduces a strong splice donor site at this position and consequently the activation of the above mentioned cryptic splice acceptor sites (**Figure2b**). Both aberrant transcripts contain a predicted premature stop codon and are consequently expected to result in loss-of-function through either nonsense-mediated mRNA decay (NMD), or an abnormal protein product (**Figure 2b**). Comparing various heterozygous single nucleotide variants shows modest allelic imbalance in the RNA sequence versus DNA, which indicates some NMD (**Supplemental Figure 6)**. Of interest, this variant was described before, with similar effects on RNA splicing characterized by RT-PCR.^35^

**Figure 2.**
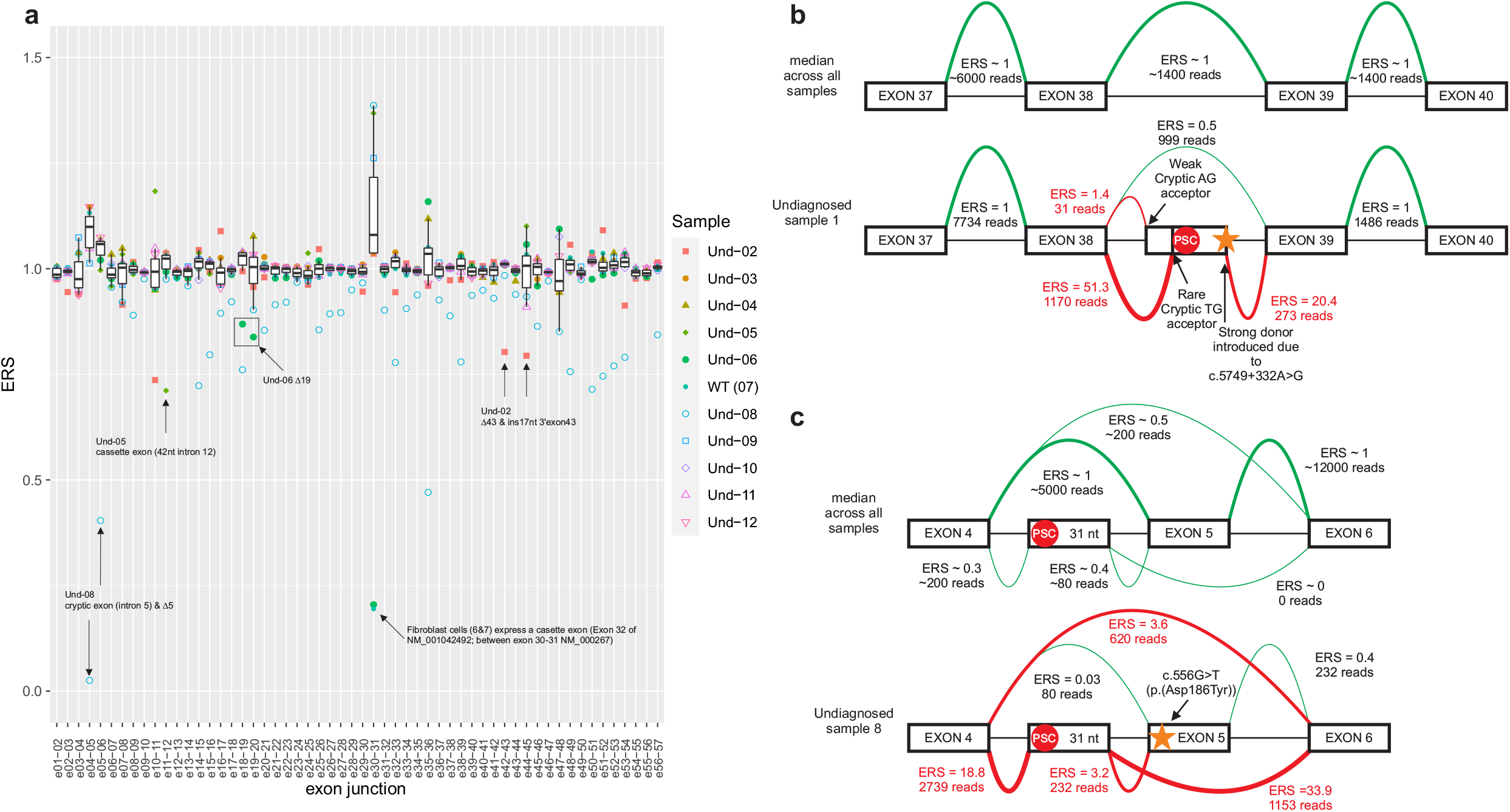
For all patient derived samples with a pathogenic splicing event a concomitant drop of the normal exon junction in close proximity is visible. Enrichment score of normal exon-exon splice junctions of the undetermined cohort (a), annotated in the figure are the drop in ERS due to the detected pathogenic splicing event. Normal reference exon splice junctions are expected to have a median ERS ∼1. Splicing of undetermined 1 and 8 explained (b/c). Shown are the median ERS and reads for all samples within the same run vs the actual ERS and reads of the sample indicated; undiagnosed 1 (b) and undiagnosed 8 (c).

Patient 2 was included with a likely pathogenic (class 4) variant c.6580-2A>G, which is predicted to show loss of the acceptor splice site that could lead to exon skipping. The splicing predictions however also indicate multiple cryptic acceptor sites upstream (**Supplemental Figure 7**) that could be alternatively used. Experimental analysis showed exon 43 skipping (ERS 8.5, 136 reads) and insertion of 17nt intron sequence through use of the first upstream cryptic acceptor (ERS 7.1, 97 reads) was observed (**Supplemental Figure 7**), leading to a drop in ERS of the normal exon-exon junction (**Figure 2a)**. The variant is visible in the RNAseq reads (**Supplemental Figure 7**). No allelic imbalance was observed (data not shown) despite absence of an NMD inhibitor in the blood samples, suggesting that the truncated protein is synthesized and not degraded at the mRNA level by NMD. Based on these findings the c.6580-2A>G variant is now considered pathogenic (class 5).

For patient 5, a cassette exon including 42 bp of intron 11 was detected (acceptor site ERS 18.1 and 1497 reads, donor site ERS 28 and 2278 reads), which introduces a premature stop codon. (**Supplemental Figure 8)**. This event was found based on high ERS (**Table 3, Supplemental Table 5)** and a drop in ERS of the normal exon-11-12 junction (**Figure 2a**). The aberrant transcript is caused by a c.1260+1604A>G change that is visible in the RNAseq reads (**Supplemental Figure 8)** and confirmed with Sanger DNA sequence analysis. Comparing various single nucleotide variants shows modest allelic imbalance in the RNA sequence versus DNA sequencing reads, which indicates some NMD (**Supplemental Figure 8)**. Interestingly, the pathogenic splicing variant c.1260+1604A>G is reported multiple times in the literature and found to have a similar effect on splicing.^36,37^

Patient 6 has a clinical diagnosis of NF1 and a variant of unknown significance (VUS) c.2252G>T (p.(Gly751Val)) in exon 19 of *NF1*. Her child also has the same variant and a clinical NF1 diagnosis. The glycine is conserved between species, the variant is not present in gnomAD and has, as annotated by Alamut strong disease causing *in silico* predictions. These predictions indicate partial loss of the (strong) acceptor site and introduction of a new donor site, potentially resulting in out-of-frame exon 19 skipping (**Supplemental Figure 9**). For this patient, RNA from short term cultured fibroblast was used, without NMD inhibition. QURNAs indicated exon 19 skipping, which is a naturally occurring event, hence the ERS is only 4.2 (covered by 4155 reads). The drop in ERS of the normal exon 18-19 junction is prominent though (**Figure 2a, Supplemental Figure 9**). Of interest, comparing various single nucleotide variants, including c.2252G>T, showed almost a complete loss of one allele in the RNA sequence versus DNA sequencing reads, suggesting NMD of the variant allele (**Supplemental Figure 9**). This variant was previously also reported in other NF1-patients.^38^ Together, these data support the classification of this variant as pathogenic.

For patient 8, RNA was directly isolated from peripheral blood lymphocytes, in contrast to the other samples in this run. This patient has a clinical diagnosis of NF1 and a VUS (class 3 variant) c.556G>T (p.(Asp186Tyr)) in exon 5 of *NF1*. The aspartic acid is conserved between species and the variant is not present in gnomAD, as well as having strong disease causing *in silico* predictions (as annotated by Alamut). Splicing predictions show no effect on splicing through the introduction of a novel donor or acceptor site. Only a minor effect on an exonic splicing enhancer element and a slightly higher chance of exon skipping than the wild type (WT) allele was predicted (**Supplemental Figure 10**). Interestingly, literature suggests that the variant c.557A>T immediately next to the one identified in this patient (c.556G>T) is disrupting an exonic splicing enhancer element, thereby causing exon skipping.^39^ Therefore this sample was included in this RNAseq study and found indeed to cause some skipping of exon 5 (ERS 3.6; 620 reads), but inclusion of a cryptic intron between exons 4 and 5 (acceptor ERS 33.9 and 1153 reads and donor ERS 18.8 and 2739 reads) is the major event (**Figure 2b)**. Both events are naturally occurring albeit that the drop in ERS of exon boundaries 4-5 and 5-6 for this patient is more prominent compared to the others in the same run (**Figure 2a**). Of interest, comparing the RNA versus the DNA sequencing reads showed partial loss of the alternative T allele of the c.556G>T variant, caused by skipping of this exon. Some, but not all, of the other single nucleotide variants indicated weak allelic imbalance (**Supplemental Figure 10**), which suggests that the premature stop does not subject the transcript to strong NMD. Together, these data support the classification of this variant as likely pathogenic. Further segregation analysis should be performed to support pathogenicity.

## Discussion

Alternative splicing is an important mechanism wherein different mRNAs are generated from the same gene, thus increasing the coding capacity. It is often regulated in a tissue- or developmental-specific manner. Deregulation of this mechanism can be caused by DNA variants in regulatory regions of RNA splicing, resulting in aberrant splicing. Skipping of exons and/or the introduction of a premature stop codon can lead to loss-of-function or even complete loss of the encoded protein from the variant allele. Several naturally occurring alternative transcripts have been described for the *NF1* gene as well as aberrant splicing events due to DNA variants in this gene. In fact, a high percentage of up to ∼30% of pathogenic *NF1* DNA variants have an effect on RNA splicing. Targeted RNA sequencing is currently more cost efficient compared to whole transcriptome for in depth analysis of the RNA splicing of a small set of genes. Here we show that targeted *NF1* RNAseq, in combination with QURNAS data-analysis (**Supplemental Figure 1**), is a powerful approach to detect and distinguish normal and aberrant, pathogenic *NF1* splicing events.

Not only could targeted RNAseq confirm previously reported and/or predicted changes in RNA splicing for all validation and replication samples, additional splicing events caused by the same DNA variant, but not (easily) resolved using RT-PCR, could be detected (**Table 1,2**). In these samples, with RNA extracted from lymphocytes, all normal exon-exon junctions and rare normal events previously reported in blood derived RNA could also be detected, except the neuronal tissue specific inclusion of a cassette exon of 30nt intron 11 (‘ins 9a/9br’). Additionally, the use of two different enrichment methods gave similar results (**Table 2, Supplemental Figure 5**).

After this thorough validation of our targeted RNAseq approach a total of 11 patients lacking a (clear) molecular diagnosis of NF1 was tested. Three patients had a clinical NF1 diagnosis and a *NF1* DNA variant suspected to have an effect on *NF1* splicing. The other eight patients showed some or clear NF1 symptoms, but lacked a molecular diagnosis through conventional DNA sequencing of *NF1* (and *SPRED1)*. Pathogenic alternative *NF1* splicing could be detected in five out of these 11 patients using RNAseq (45%). For three of them this was a confirmation of the effect suspected by *in silico* splice predictions for a DNA variant detected with prior DNA analysis. For two patients (of eight without a prior molecular diagnosis) the pathogenic splicing event was caused by a deep-intronic DNA variant (25%), that was previously not detected using standard DNA diagnostics.

For six patients, no aberrant, pathogenic *NF1*, nor *SPRED1* RNA splicing or expression could be detected. In most cases, bi-allelic gene expression could be confirmed, excluding the presence of a possible variant in the promoter region or transcription enhancer site that could result in strongly reduced or absent gene expression in the variant allele. To exclude other potential (rare) causes of NF1, the 5’UTR (up to c.-383) and the neuronal specific cassette exon of 30nt in intron 11 (‘ins 9a/9br’) of *NF1* were also sequenced complementary, since these regions are not covered by RNAseq, nor by our standard DNA diagnostics procedure. Variants in the 5’UTR of *NF1* creating or removing an upstream start codon (uAUG) and thus influencing translation can cause NF1.^14,40^ No relevant variants were however detected in these 2 regions.

In addition, retrotransposon insertions, that cause altered NF1 splicing, account for ∼0.4% of all pathogenic *NF1* variants.^17^ These are not targeted using conventional DNA diagnostics and not detected using RNAseq due to the exclusion of chimeric reads in in the RNAseq data analysis pipeline. Another residual risk of missing variants in blood based DNA and RNA diagnostics is the presence of a mosaic postzygotic *NF1* pathogenic variant, in specific affected tissue, but absent in blood. This might be the case for patient 4 (Table 3) who presented with a segmental/mosaic NF1 phenotype restricted to the skin. Unfortunately, this affected tissue was not available for further DNA or RNA analysis. Finally, an NF1 (like) clinical phenotype, might also be explained by molecular defects in other genes, causing syndromes with overlapping phenotypes. These are for example Noonan syndrome (MIM PS163950; multiple genes/loci), Constitutional mismatch repair deficiency (MIM 276300; mismatch repair genes) and Proteus syndrome (MIM 176920; *AKT1*).^1,2,5^

The QURNAs algorithm facilitated the identification of significant changes in RNA splicing. It uses an unbiased approach to identify and characterize aberrant and novel splice junctions without prior assumptions about transcript isoforms. The pipeline identifies all splice events present in the data and calculates their enrichment score and statistical significance compared to other samples within the same run. Together with a generic targeted RNAseq procedure, the QURNAs method can be used in routine diagnostics complementary to DNA diagnostics to discover, confirm or exclude effect of genetic variants on RNA splicing in NF1 or other genes of interest.

In conclusion, the presented targeted RNAseq approach in combination with the computational QURNAs pipeline successfully detects and quantifies pathogenic *NF1* splicing events driven by (deep) intronic or missense variants, complementary to DNA diagnostics.

## Data Availability

Data is available upon request

## Acknowledgements/Author information

We thank Roel Brandts for excellent technical assistance in *NF1* DNA and RNA diagnostics.

## Notes

The authors declare no conflicts of interest.

### Competing Interest Statement

The authors have declared no competing interest.

### Funding Statement

no external funding

### Author Declarations

We obtainend an excemption from the 'medisch-ethische toetsingscommissie (METC)' (translated medical ethics review committee) of the university hospital and university of Maastricht. METC 2021-2666 stating: "To whom it may concern: we are pleased to confirm that the Medical Research Involving Human Subjects Act (WMO) does not apply to the above mentioned study and that an official approval of this study by our committee is not required."

## References

1. Friedman JM. Neurofibromatosis 1. In: Adam MP, Ardinger HH, Pagon RA, et al., editors. GeneReviews((R)). Seattle (WA)1993.

2. Ferner RE, Huson SM, Thomas N, et al. Guidelines for the diagnosis and management of individuals with neurofibromatosis 1. J Med Genet. 2007;44(2):81–88.

3. Friedman JM, Gutmann DH, MacCollin M, Riccardi VM, eds. Neurofibromatosis: Phenotype, Natural History and Pathogenesis 3rd ed. Baltimore, MD: The Johns Hopkins University Press; 1999.

4. Health NIo. National Institutes of Health Consensus Development Conference Statement: neurofibromatosis. Bethesda, Md., USA, July 13-15, 1987. Neurofibromatosis. 1988;1(3):172–178.

5. Perez-Valencia JA, Gallon R, Chen Y, et al. Constitutional mismatch repair deficiency is the diagnosis in 0.41% of pathogenic NF1/SPRED1 variant negative children suspected of sporadic neurofibromatosis type 1. Genet Med. 2020.

6. Cawthon RM, O’Connell P, Buchberg AM, et al. Identification and characterization of transcripts from the neurofibromatosis 1 region: the sequence and genomic structure of EVI2 and mapping of other transcripts. Genomics. 1990;7(4):555–565.

7. Viskochil D, Buchberg AM, Xu G, et al. Deletions and a translocation interrupt a cloned gene at the neurofibromatosis type 1 locus. Cell. 1990;62(1):187–192.

8. Wallace MR, Marchuk DA, Andersen LB, et al. Type 1 neurofibromatosis gene: identification of a large transcript disrupted in three NF1 patients. Science. 1990;249(4965):181–186.

9. Trovo-Marqui AB, Tajara EH. Neurofibromin: a general outlook. Clin Genet. 2006;70(1):1–13.

10. Stenson PD, Mort M, Ball EV, et al. The Human Gene Mutation Database: towards a comprehensive repository of inherited mutation data for medical research, genetic diagnosis and next-generation sequencing studies. Hum Genet. 2017;136(6):665–677.

11. Landrum MJ, Lee JM, Benson M, et al. ClinVar: improving access to variant interpretations and supporting evidence. Nucleic Acids Res. 2018;46(D1):D1062–D1067.

12. van Minkelen R, van Bever Y, Kromosoeto JN, et al. A clinical and genetic overview of 18 years neurofibromatosis type 1 molecular diagnostics in the Netherlands. Clin Genet. 2014;85(4):318–327.

13. Wimmer K, Roca X, Beiglbock H, et al. Extensive in silico analysis of NF1 splicing defects uncovers determinants for splicing outcome upon 5’ splice-site disruption. Hum Mutat. 2007;28(6):599–612.

14. Evans DG, Bowers N, Burkitt-Wright E, et al. Comprehensive RNA Analysis of the NF1 Gene in Classically Affected NF1 Affected Individuals Meeting NIH Criteria has High Sensitivity and Mutation Negative Testing is Reassuring in Isolated Cases With Pigmentary Features Only. EBioMedicine. 2016;7:212–220.

15. Messiaen LM, Callens T, Mortier G, et al. Exhaustive mutation analysis of the NF1 gene allows identification of 95% of mutations and reveals a high frequency of unusual splicing defects. Hum Mutat. 2000;15(6):541–555.

16. Valero MC, Martin Y, Hernandez-Imaz E, et al. A highly sensitive genetic protocol to detect NF1 mutations. J Mol Diagn. 2011;13(2):113–122.

17. Wimmer K, Callens T, Wernstedt A, Messiaen L. The NF1 gene contains hotspots for L1 endonuclease-dependent de novo insertion. PLoS Genet. 2011;7(11):e1002371.

18. Richards S, Aziz N, Bale S, et al. Standards and guidelines for the interpretation of sequence variants: a joint consensus recommendation of the American College of Medical Genetics and Genomics and the Association for Molecular Pathology. Genet Med. 2015;17(5):405–424.

19. Vreeswijk M, Kraan J, Klift Hvd, et al. Intronic variants in BRCA1 and BRCA2 that affect RNA splicing can be reliably selected by splice-site prediction programs. Hum Mutat. 2008;30(1):107–114.

20. Brandão RD, van Roozendaal K, Tserpelis D, Gomez Garcia E, Blok MJ. Characterisation of unclassified variants in the BRCA1/2 genes with a putative effect on splicing. Breast cancer research and treatment. 2011;129(3):971-982 (Research Support, Non-U.S. Gov’t).

21. Sangermano R, Khan M, Cornelis SS, et al. ABCA4 midigenes reveal the full splice spectrum of all reported noncanonical splice site variants in Stargardt disease. Genome Res. 2018;28(1):100–110.

22. Houdayer C, Caux-Moncoutier V, Krieger S, et al. Guidelines for splicing analysis in molecular diagnosis derived from a set of 327 combined in silico/in vitro studies on BRCA1 and BRCA2 variants. Human Mutation. 2012;33(8):1228–1238.

23. Taggart AJ, DeSimone AM, Shih JS, Filloux ME, Fairbrother WG. Large-scale mapping of branchpoints in human pre-mRNA transcripts in vivo. Nat Struct Mol Biol. 2012;19(7):719–721.

24. Andreutti-Zaugg C, Scott RJ, Iggo R. Inhibition of nonsense-mediated messenger RNA decay in clinical samples facilitates detection of human MSH2 mutations with an in vivo fusion protein assay and conventional techniques. Cancer Res. 1997;57(15):3288–3293.

25. Dobin A, Davis CA, Schlesinger F, et al. STAR: ultrafast universal RNA-seq aligner. Bioinformatics. 2013;29(1):15–21.

26. den Dunnen JT, Antonarakis SE. Mutation nomenclature extensions and suggestions to describe complex mutations: a discussion. Hum Mutat. 2000;15(1):7–12.

27. Danglot G, Regnier V, Fauvet D, Vassal G, Kujas M, Bernheim A. Neurofibromatosis 1 (NF1) mRNAs expressed in the central nervous system are differentially spliced in the 5’ part of the gene. Hum Mol Genet. 1995;4(5):915–920.

28. Gutmann DH, Zhang Y, Hirbe A. Developmental regulation of a neuron-specific neurofibromatosis 1 isoform. Ann Neurol. 1999;46(5):777–782.

29. Geist RT, Gutmann DH. Expression of a developmentally-regulated neuron-specific isoform of the neurofibromatosis 1 (NF1) gene. Neurosci Lett. 1996;211(2):85–88.

30. Ars E, Serra E, Garcia J, et al. Mutations affecting mRNA splicing are the most common molecular defects in patients with neurofibromatosis type 1. Hum Mol Genet. 2000;9(2):237–247.

31. Pros E, Gomez C, Martin T, Fabregas P, Serra E, Lazaro C. Nature and mRNA effect of 282 different NF1 point mutations: focus on splicing alterations. Hum Mutat. 2008;29(9):E173–193.

32. De Conti L, Skoko N, Buratti E, Baralle M. Complexities of 5’splice site definition: implications in clinical analyses. RNA Biol. 2012;9(6):911–923.

33. Vandenbroucke I, Vandesompele J, De Paepe A, Messiaen L. Quantification of NF1 transcripts reveals novel highly expressed splice variants. FEBS Lett. 2002;522(1-3):71–76.

34. Messiaen LM, Callens T, Roux KJ, et al. Exon 10b of the NF1 gene represents a mutational hotspot and harbors a recurrent missense mutation Y489C associated with aberrant splicing. Genet Med. 1999;1(6):248–253.

35. Perrin G, Morris MA, Antonarakis SE, Boltshauser E, Hutter P. Two novel mutations affecting mRNA splicing of the neurofibromatosis type 1 (NF1) gene. Hum Mutat. 1996;7(2):172–175.

36. Sabbagh A, Pasmant E, Imbard A, et al. NF1 molecular characterization and neurofibromatosis type I genotype-phenotype correlation: the French experience. Hum Mutat. 2013;34(11):1510–1518.

37. Serra E, Rosenbaum T, Winner U, et al. Schwann cells harbor the somatic NF1 mutation in neurofibromas: evidence of two different Schwann cell subpopulations. Hum Mol Genet. 2000;9(20):3055–3064.

38. Stella A, Lastella P, Loconte DC, et al. Accurate Classification of NF1 Gene Variants in 84 Italian Patients with Neurofibromatosis Type 1. Genes (Basel). 2018;9(4).

39. Zatkova A, Messiaen L, Vandenbroucke I, et al. Disruption of exonic splicing enhancer elements is the principal cause of exon skipping associated with seven nonsense or missense alleles of NF1. Hum Mutat. 2004;24(6):491–501.

40. Whiffin N, Karczewski KJ, Zhang X, et al. Characterising the loss-of-function impact of 5’ untranslated region variants in 15,708 individuals. Nat Commun. 2020;11(1):2523.

