## Supplemental Figures for "Pathogenic Neurofibromatosis type 1 (NF1) RNA splicing resolved by targeted RNAseq"

**Supplemental Figure 1.** Targeted RNAseq-based analysis to detect normal and pathogenic *NF1* RNA-splicing

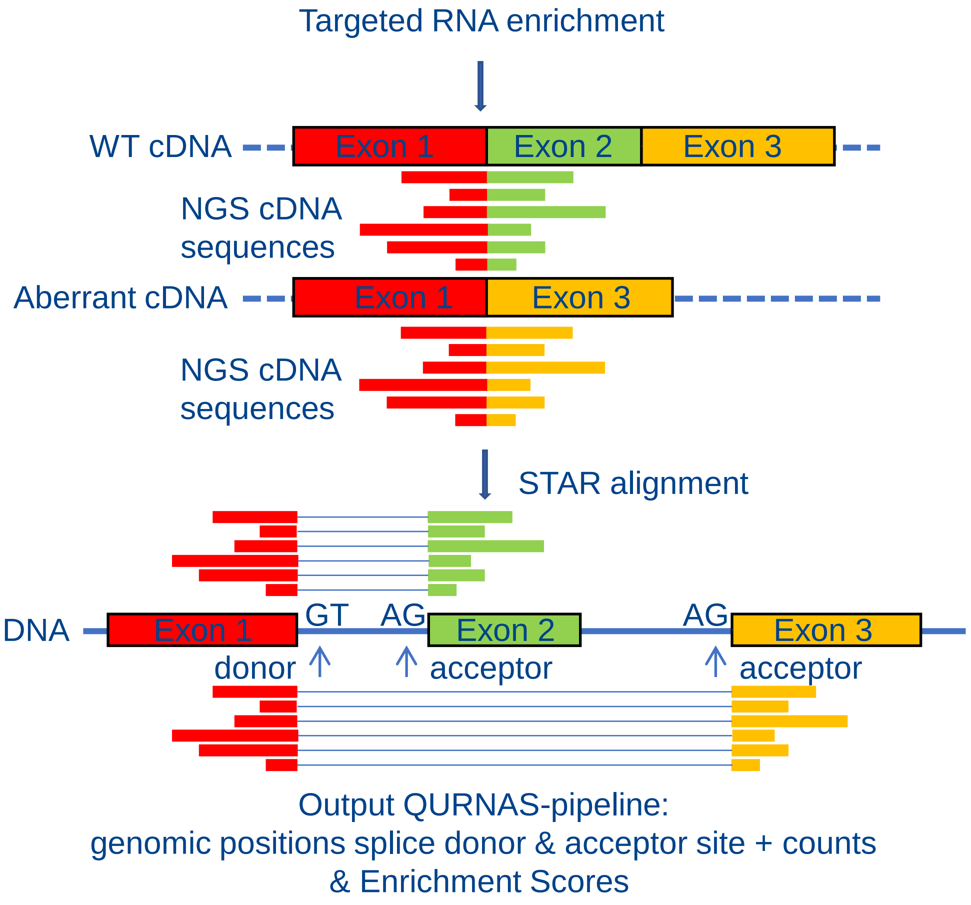


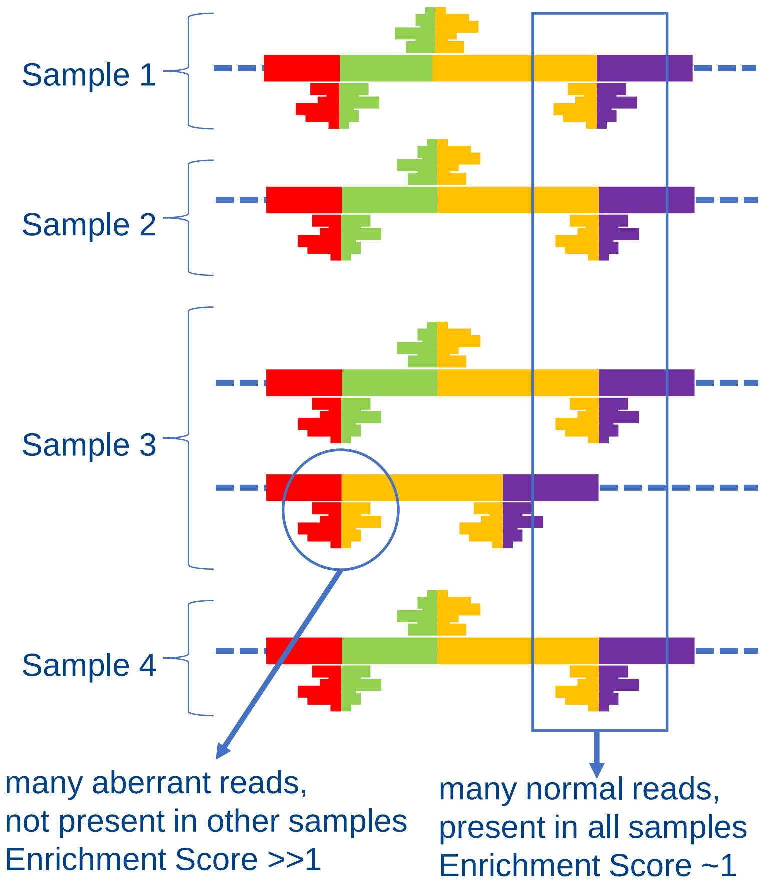


| Genomic position | | Enrichment Score (ERS) | | | | |
|---|---|---|---|---|---|---|
| Donor site | Acceptor site | Sample 1 | Sample 2 | Sample 3 | Sample 4 | |
| 29527612 | 29528054 | 1.0 | 1.2 | **0.6** | 0.9 | Exon 1-2 junction |
| 29527612 | 29528428 | 0 | 0 | **18** | 0 | Exon 2 skip |
| 29528176 | 29528428 | 0.9 | 1.1 | **0.7** | 1.0 | Exon 2-3 junction |
| 29528502 | 29533257 | 1.1 | 1.0 | 1.0 | 0.9 | Exon 3-4 junction |

**Supplemental Figure 2.** Top QURNAS events for validation sample 4 and 5 (both WT-NF1) are artefacts.

**Validation sample 4** (WT)

Highest event: deletion (Δ) 5'exon 37 - 5’exon 41, c. 4986_6506del; ERS = 5; 150 reads

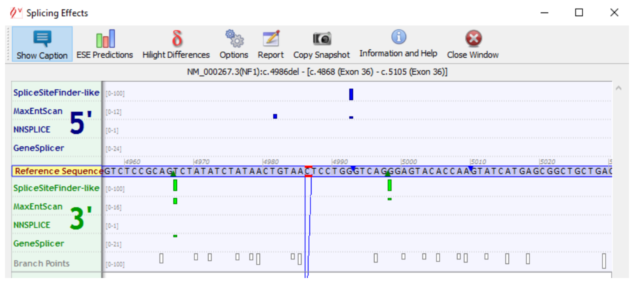


Exon 37, supposed splicing start, no *in silico* predictions for a splice donor site at this position

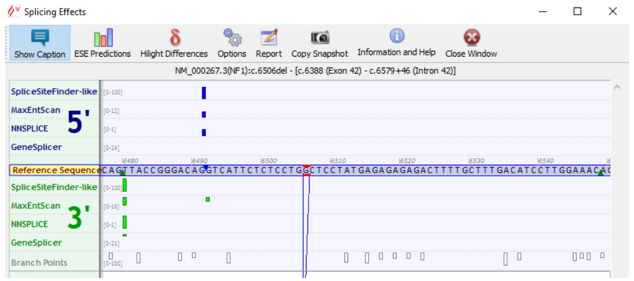


Exon 41, supposed splicing stop , no *in silico* predictions for a splice acceptor site at this position

No drop in ERS of the normal exon junctions for Val-04 (WT)

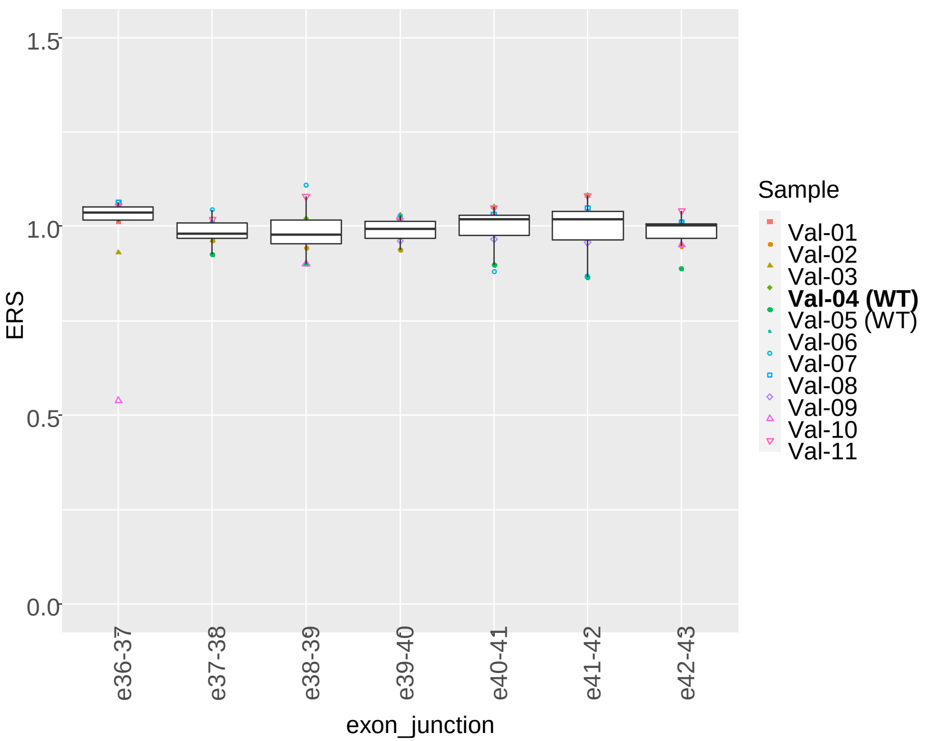


**Validation sample 5** (WT)

Δ 3’UTR c.-85_-68del; ERS 6.7, 90 reads

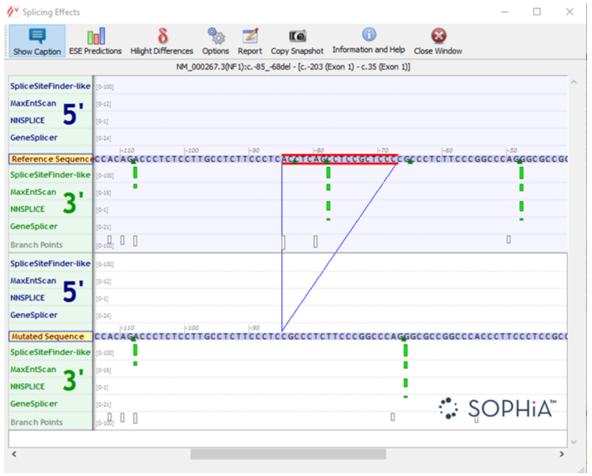


3’UTR, supposed splicing start/stop , no *in silico* predictions for a splice donor/acceptor at these positions => small deletion/artefact

### **Supplemental Figure 3.** *In silico* predictions for effect c.586+5G>A (left) and observed changes in splicing for validation sample 6, versus median values across all samples in the run (right).

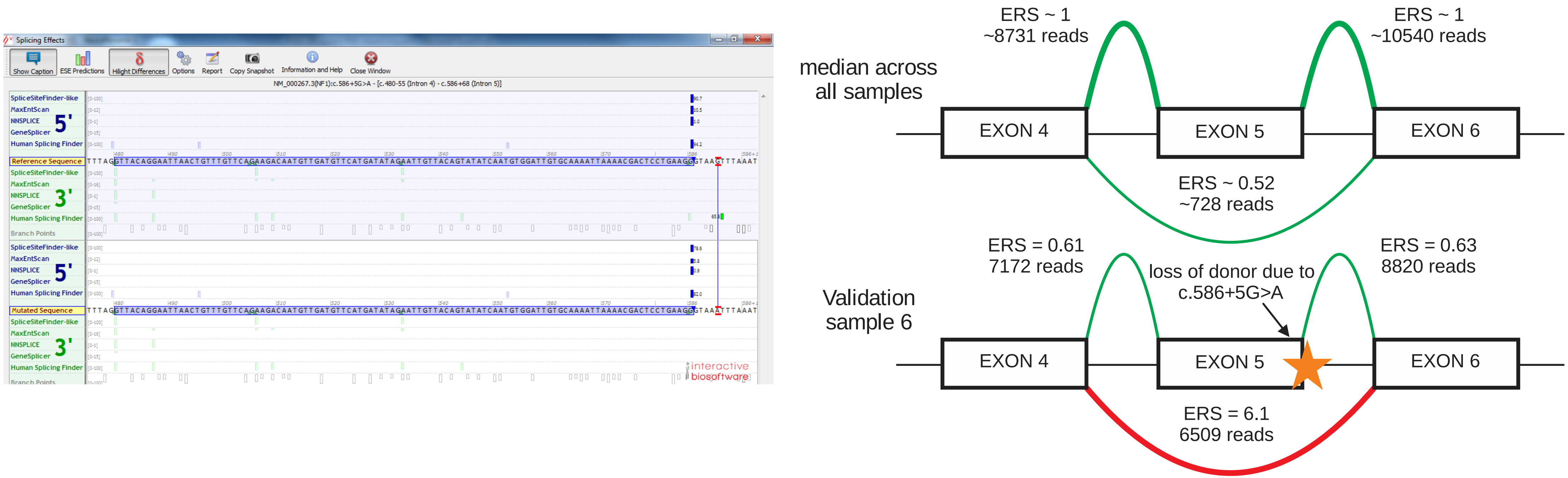

**Supplemental Figure 4.** *In silico* predictions for effect c.1466A>G (p.Tyr489Cys) and observed changes in splicing for validation sample 7 versus median values across all samples in the run (left). The QURNAS top event, Δ45nt exon 18 (IF) is an artefact (right).

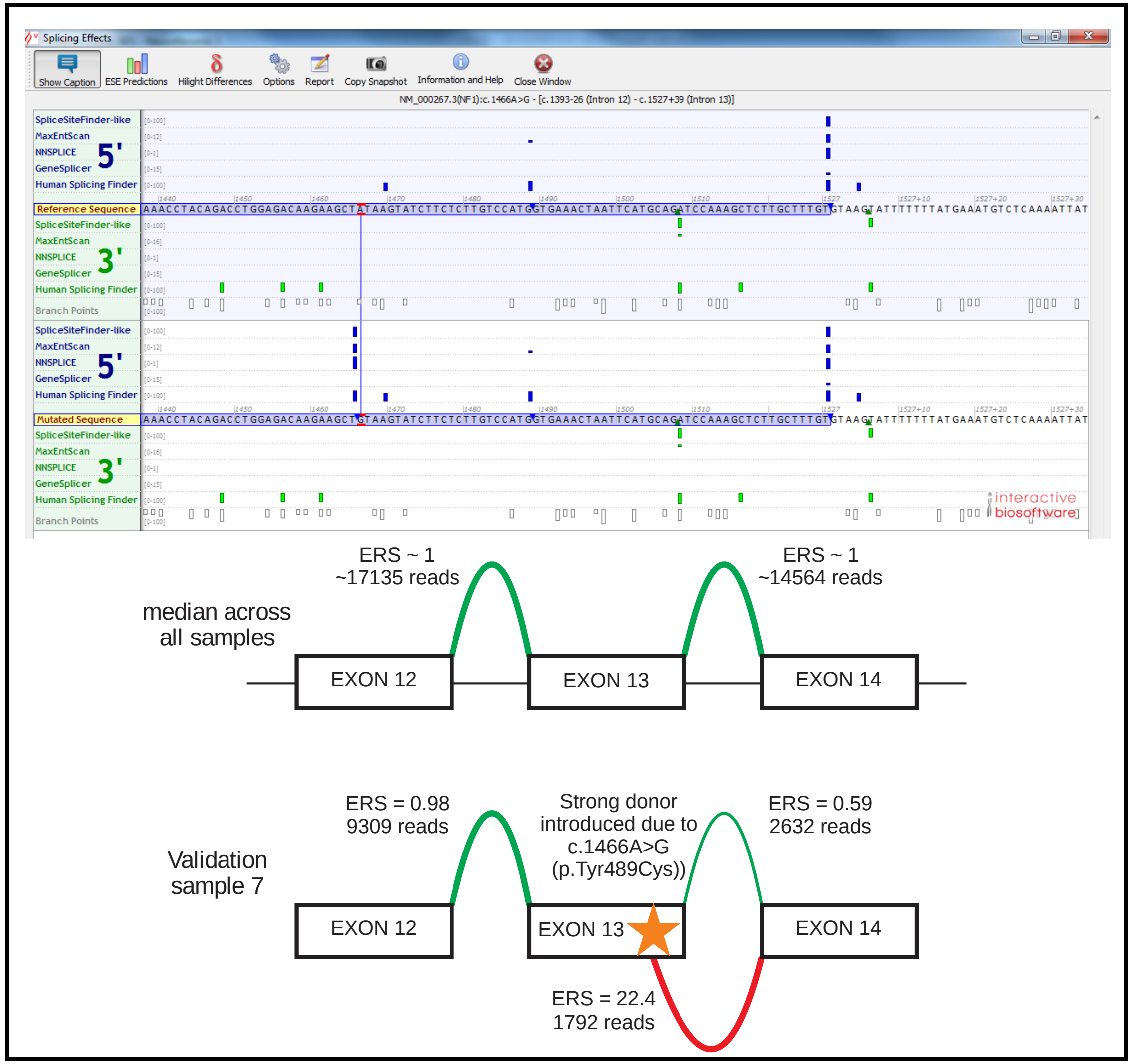


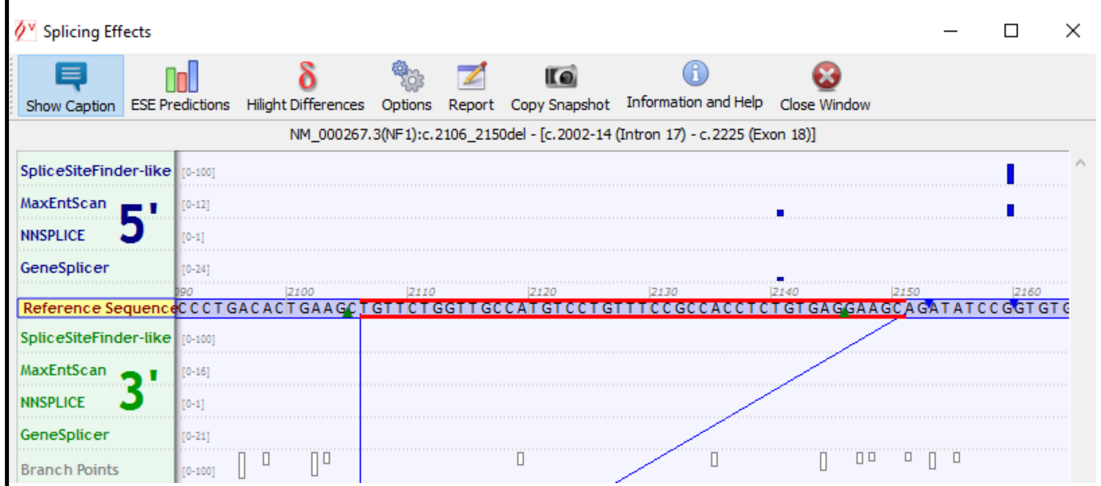


The Δ45nt are depicted in red. Note that no acceptor nor donor site are predicted that could explain the splicing of these 45nt.

**Supplemental Figure 5.** ERS values reference exon–exon splice junctions. Validation SureSelect capture using part of the replication samples.

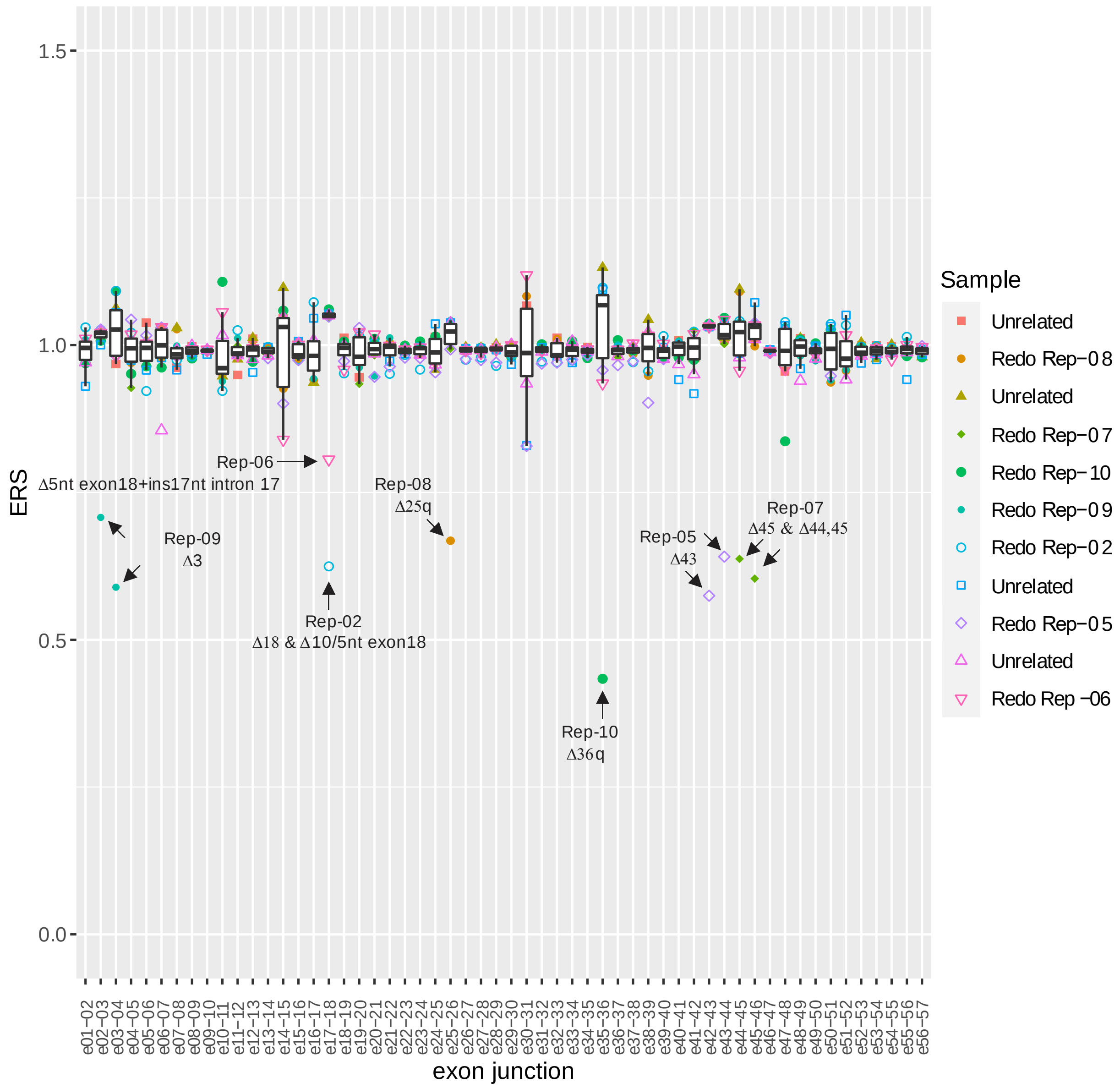

**Supplemental figure 6.** Undiagnosed sample 1 - c.5749+332A>G. This variant can be observed in the cDNA reads in a genome browser (IGV). Modest allelic imbalance allele RNA versus DNA sequence reads (left). *In silico* predictions splice effect for the c.5749+332A>G variant (right).

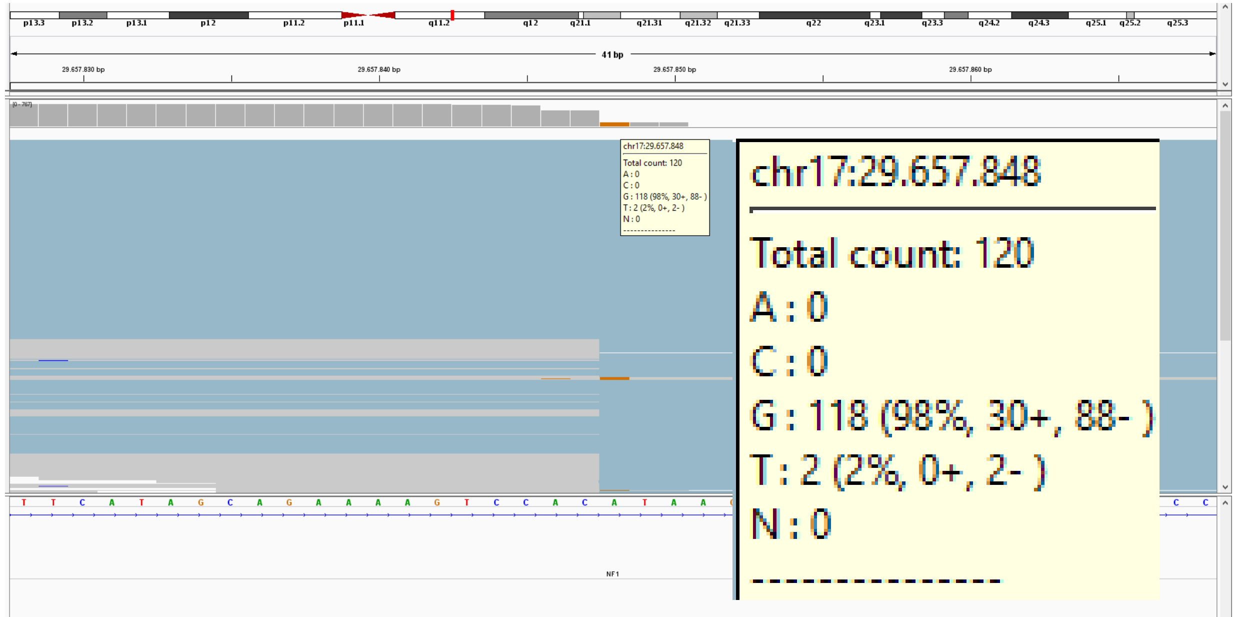


| Variant | DNA | RNA |
|---|---|---|
| c.5749+332A>G | N/A | G 98% |
| c.702G>A | A 49% | A 41% |
| c.2034G>A | A 52% | A 30% |
| c.*2265C>G | G 49% | G 31% |
| c.*2829T>C | C 51% | C 29% |

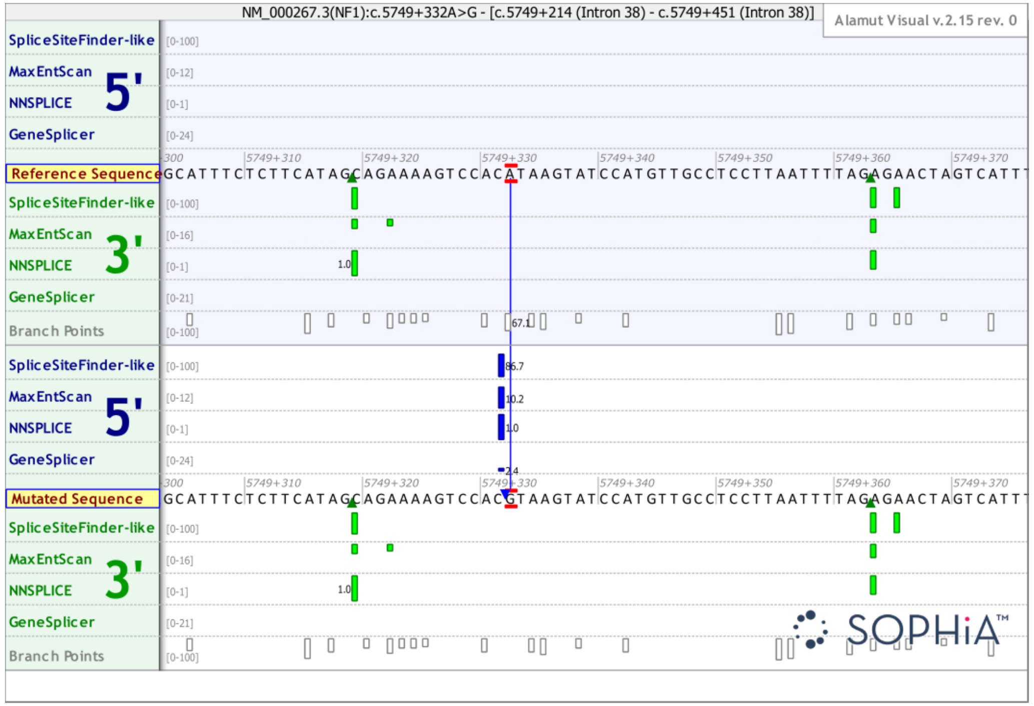

**Supplemental Figure 7.** Undiagnosed sample 2; *In silico* predictions for c.6580-2A>G (left) and observed changes in splicing for sample 2 versus median values across all samples in the run (right).

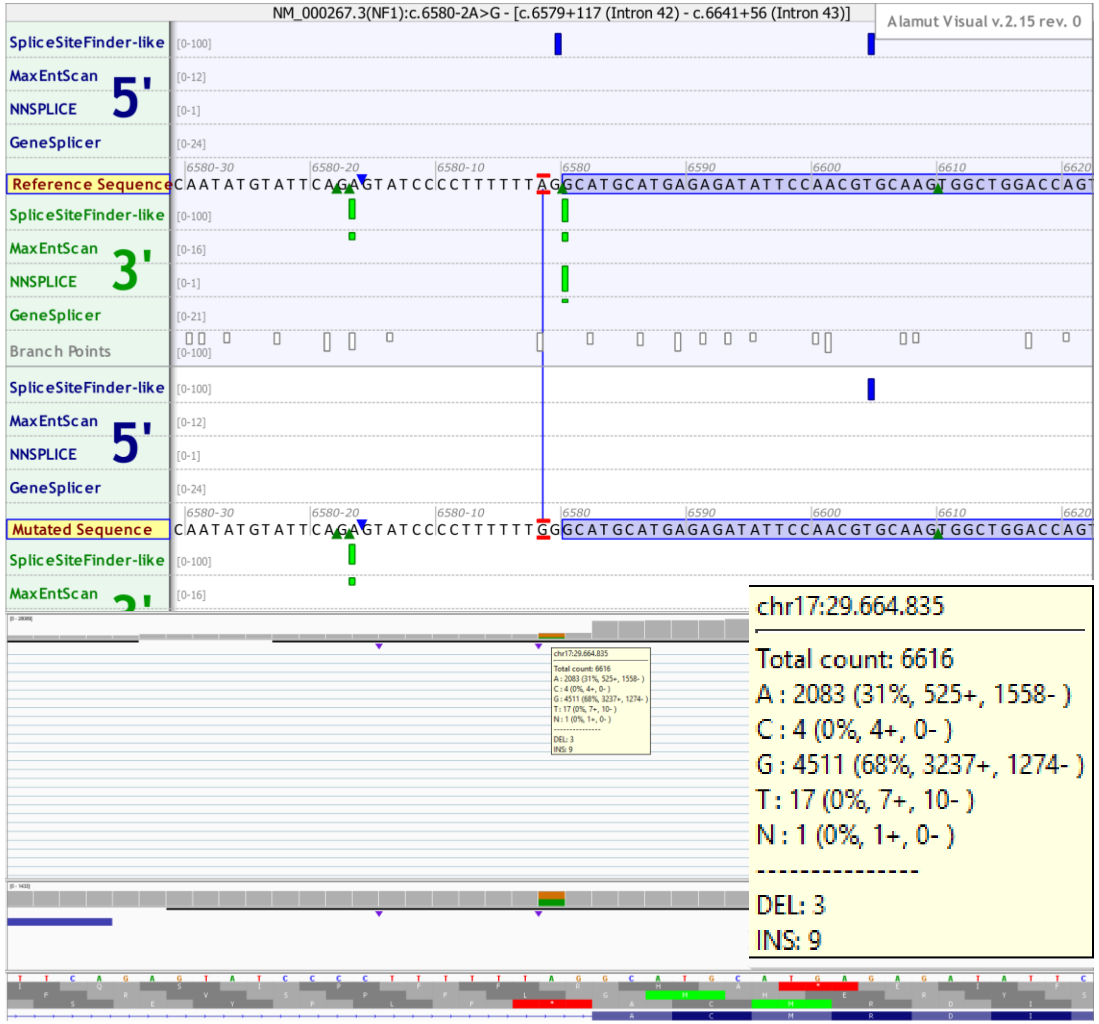


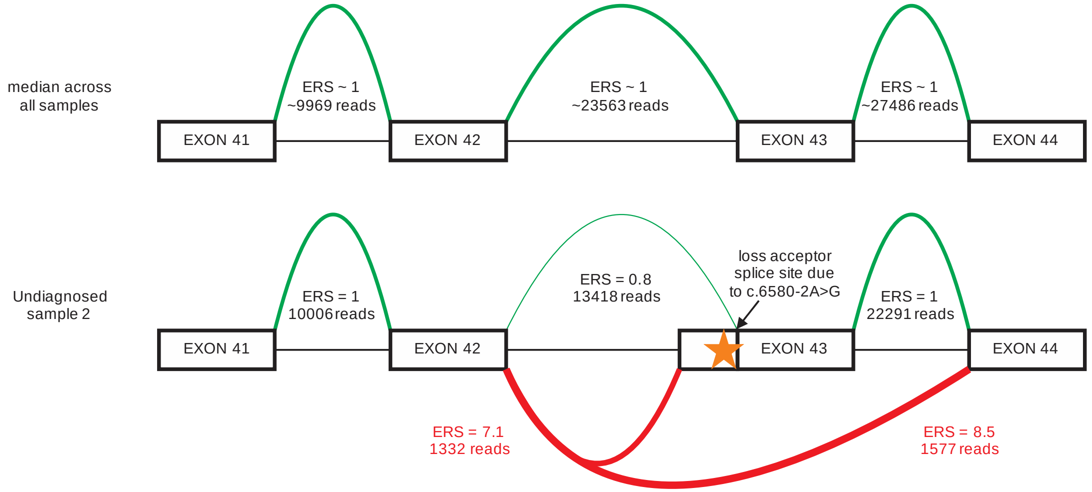

**Supplemental Figure 8.** Undiagnosed sample 5; *In silico* predictions for c.1260+1604A>G and observed changes in splicing versus median values across all samples in the run (left). Modest allelic imbalance allele RNA versus DNA sequence reads (right).

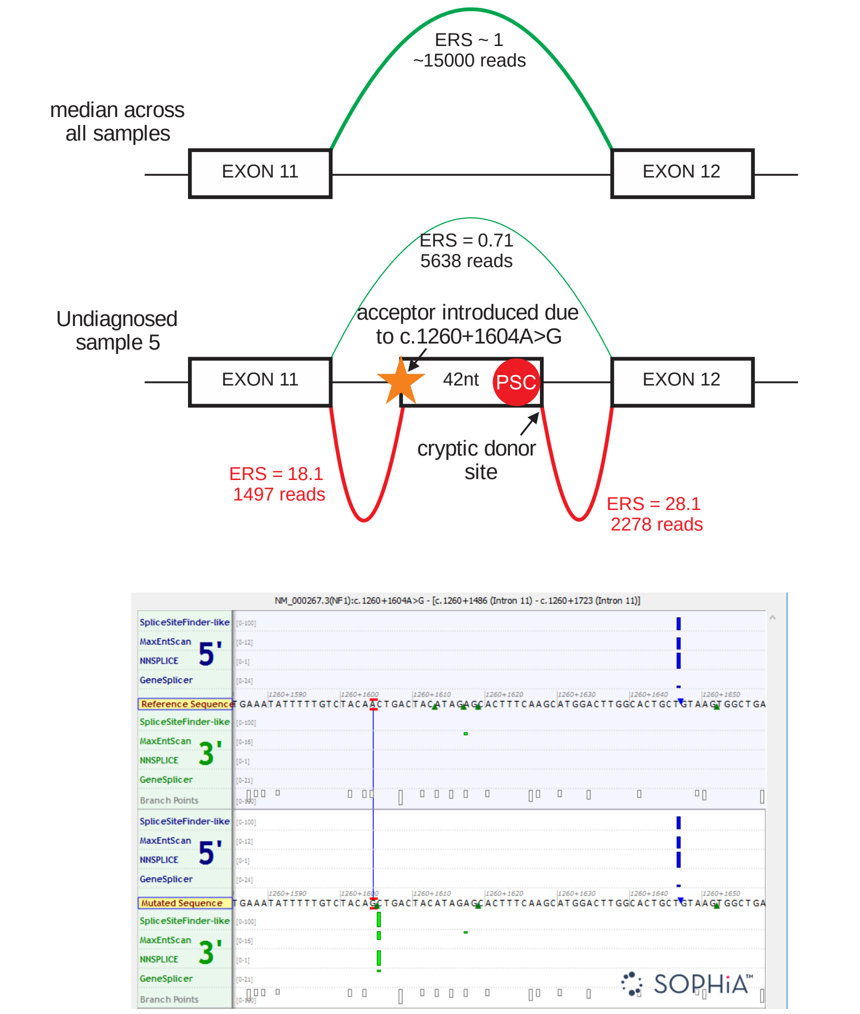


NM_000267.3(NF1):c.1260+1604A>G

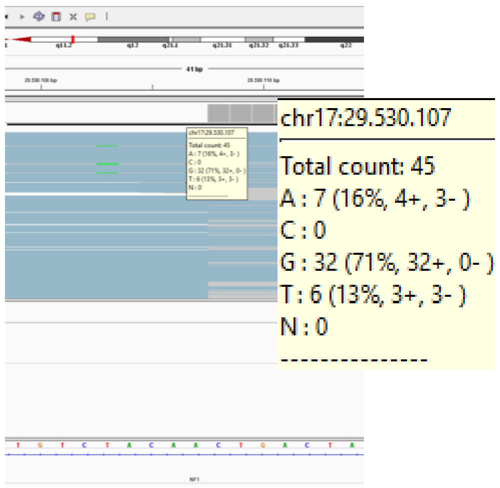


NM_000267.3(NF1):c.*2829T>C

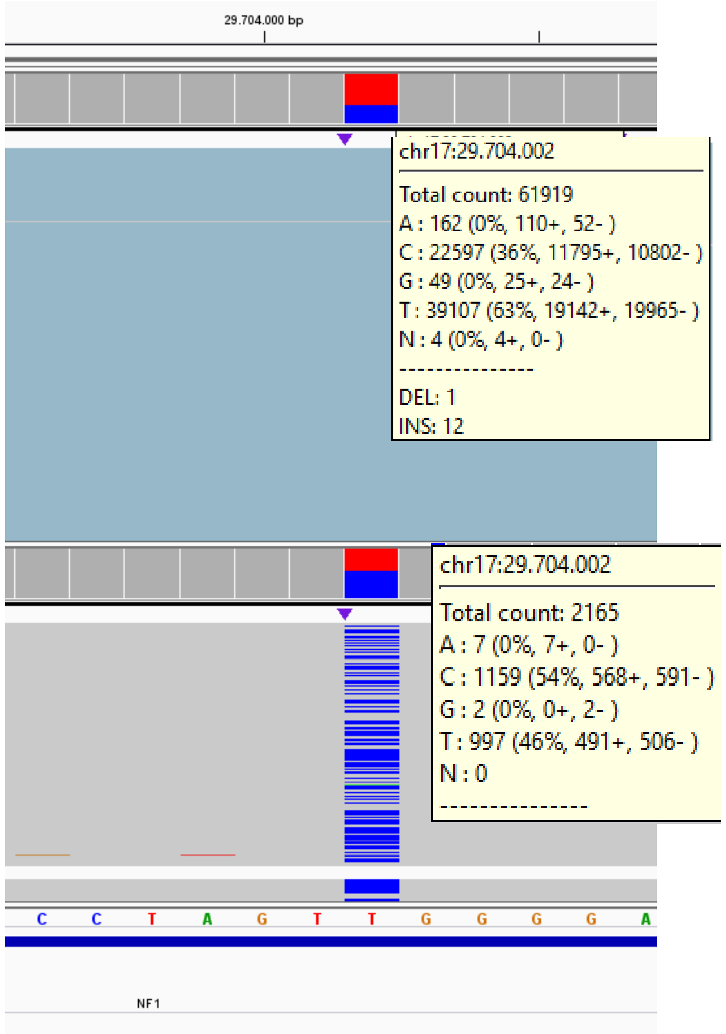


| Variant | DNA | RNA |
|---|---|---|
| c.1260+1604A>G | N/A | G 71% |
| c.168C>T | T 54% | T 35% |
| c.702G>A | A 49% | A 34% |
| c.2034G>A | A 49% | A 64% |
| c.*2829T>C | C 54% | C 36% |

**Supplemental Figure 9.** Undiagnosed sample 6; *In silico* predictions for c.2252G>T (p.(Gly751Val)) and observed changes in splicing versus median values across all samples in the run (left). Strong allelic imbalance in RNA versus DNA sequence reads (right).

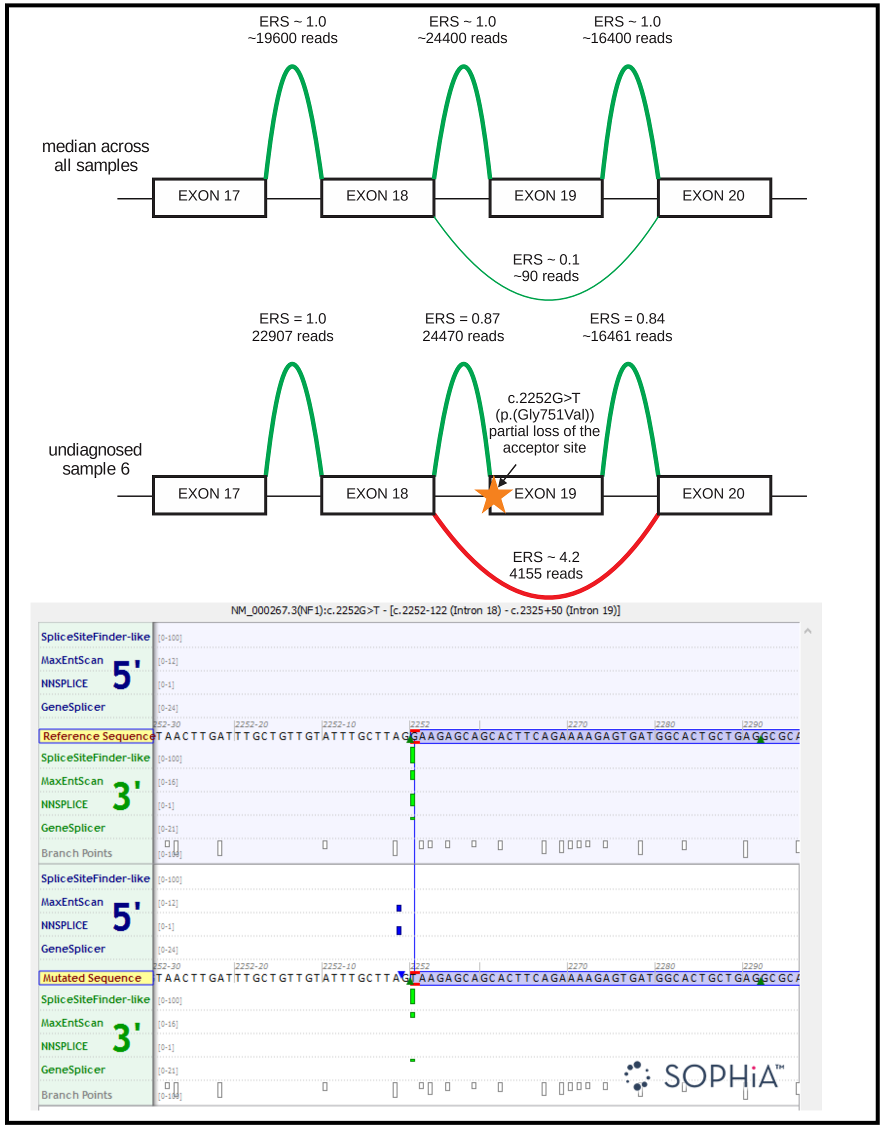


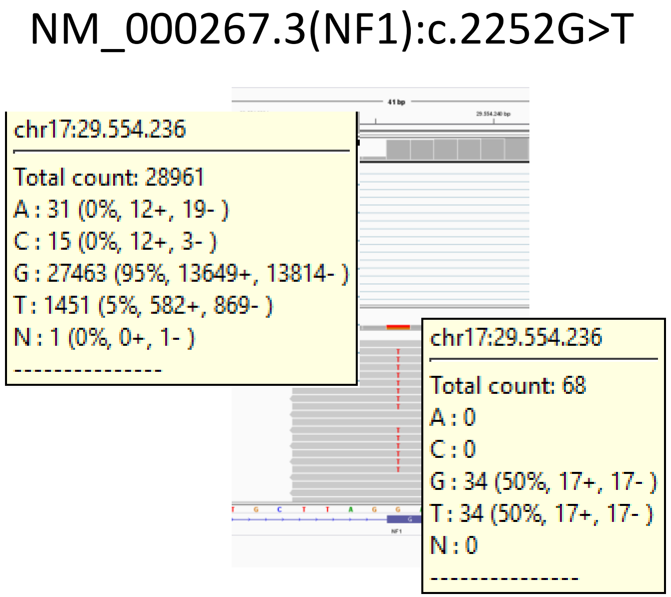


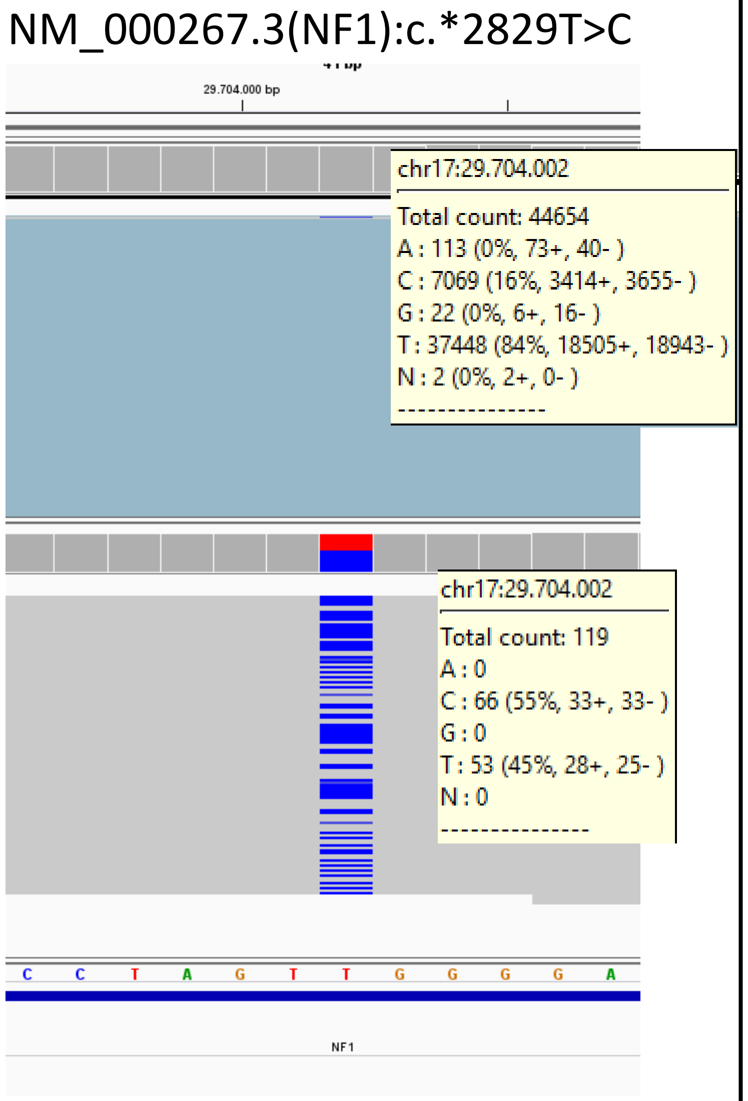


| Variant | DNA | RNA |
|---|---|---|
| c.2252G>T | T 50% | T 5% |
| c.702G>A | A 61% | A 23% |
| c.2034G>A | A 44% | A 79% |
| c.*2829T>C | C 55% | C 16% |

**Supplemental Figure 10.** Undiagnosed sample 8; *In silico* predictions for c.556G>T (p.(Asp186Tyr)) and observed changes in splicing versus median values across all samples in the run (left). Weak allelic imbalance in RNA versus DNA sequence reads (right).

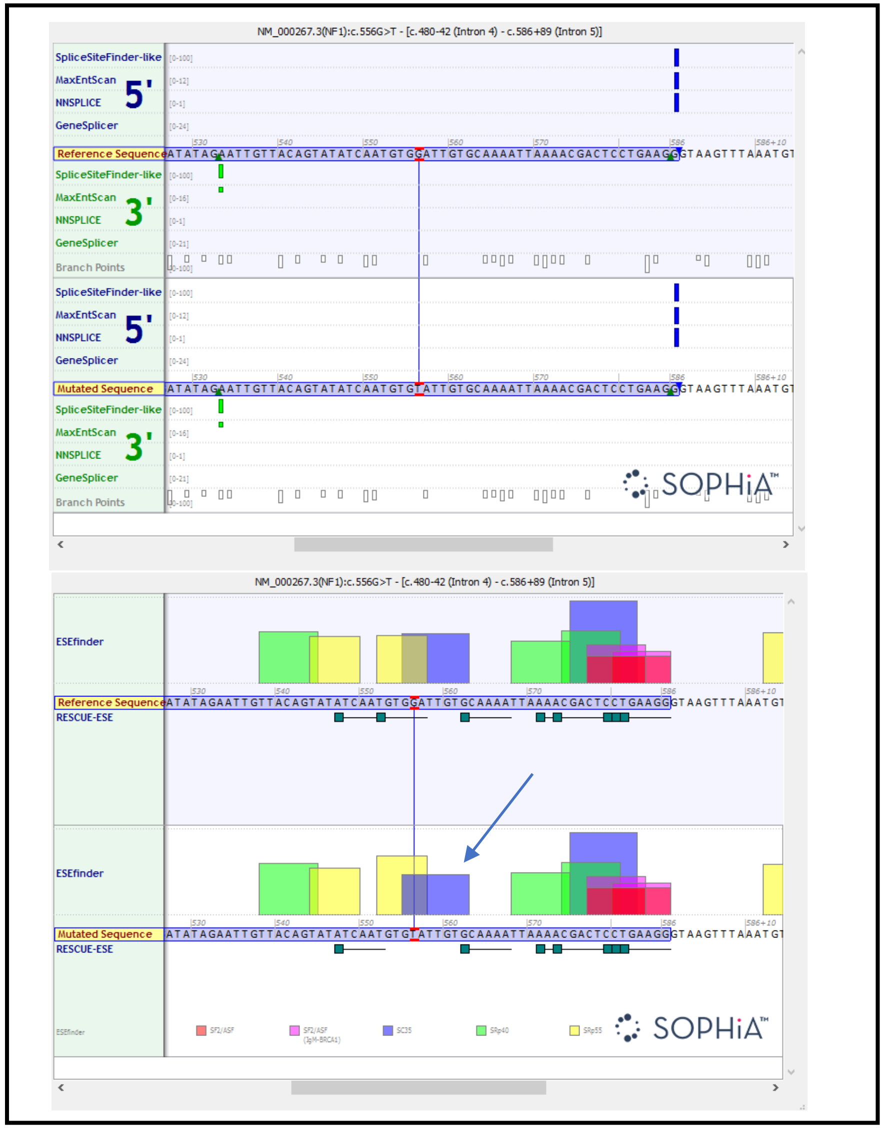


NM_000267.3(NF1):c.556G>T

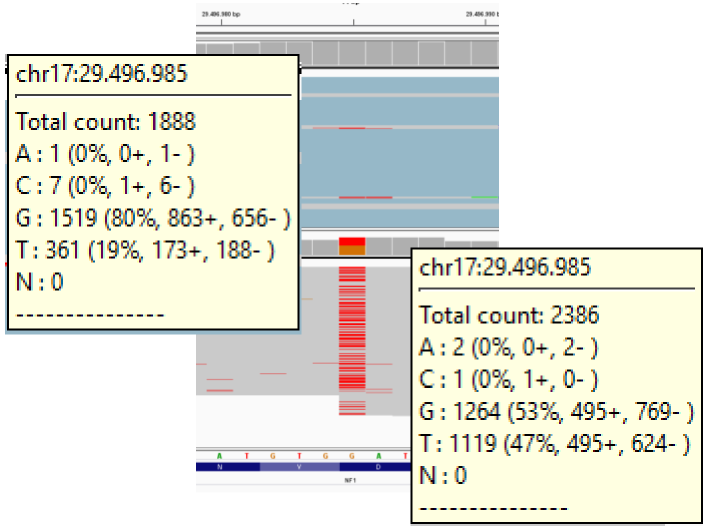


| Variant | DNA | RNA |
|---|---|---|
| c.556G>T | T 47% | T 20% |
| c.702G>A | A 48% | A 35% |
| c.2034G>A | A 50% | A 50% |
| c.*2201G>A | A 50% | A 60% |

NM_000267.3(NF1):c.*2201G>A

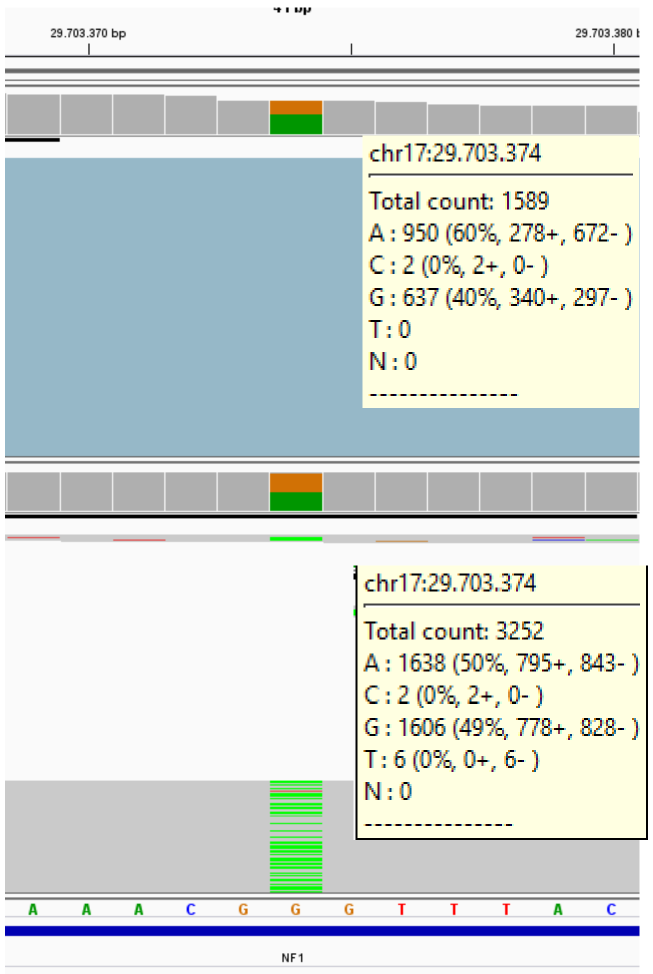
